# Anthropometric and neurocognitive consequences of *Campylobacter*, enterotoxigenic *Escherichia coli*, and norovirus: A systematic review

**DOI:** 10.1101/2025.09.10.25330400

**Authors:** Patricia Pavlinac, Gregory Zane, Ibrahim Khalil, Elizabeth Rogawski McQuade, James A. Platts-Mills, Mathias Lalika, Fatima Al-Shimari, Priyanka Shrestha, Birgitte Giersing, Mateusz Hasso-Agopsowicz

## Abstract

**Objectives:** Synthesizing the evidence of the longer-term consequences of enteric pathogens, such as stunted growth and suboptimal neurodevelopment, is a key step to articulating the value of, and generating demand for, vaccines.

**Methods:** We conducted a systematic review of published literature documenting associations of three leading causes of diarrhea (enterotoxigenic *Escherichia coli* [ETEC], norovirus, and *Campylobacter* species [sp.]) with prospective anthropometric and neurocognitive outcomes in children under five years (PROSPERO <u>CRD42024600676</u>).

**Results:** Thirty publications were included, including several reporting on data from the same underlying cohort; 16 publications included outcomes associated with *Campylobacter*, 12 ETEC, and 7 norovirus. There was large variation in how studies reported outcomes, exposure groups, and timeframes of association. There was modest evidence of linear growth detriments associated with all three pathogens, modest evidence of *Campylobacter* limiting weight gain, and no evidence of detrimental impacts of these pathogens on wasting or neurodevelopment, albeit these two outcomes were rarely reported.

**Conclusion:** Differences in outcome definitions, comparison groups, and timeframes prohibited meta-analysis and emphasize the need for more standardization of reporting anthropometric and neurocognitive outcomes following enteric pathogen infection. Randomized controlled trials of efficacious pathogen-specific interventions may help to address challenges with confounding and reverse causality in observational studies.

## Introduction

Diarrheal diseases cause over 400,000 deaths per year in children under 5 living in low and middle-income countries (LMICs)(1). Through pathways involving local and systemic inflammation and intestinal destruction, many diarrhea-causing pathogens also contribute to longer-term morbidities such as wasting, stunting, reduced school performance, and reduced earning potential, consequences which are estimated to increase deaths attributed to diarrhea by 25%.(2) Also, more than half of diarrhea episodes among children in LMICs are treated with antibiotics making diarrhea a major contributor to antimicrobial resistance (AMR).(3)

Safe and effective rotavirus vaccines are available globally and rotavirus-attributed diarrhea burden and consequences are expected to continue declining as more countries adopt rotavirus vaccines into their routine EPI schedule. Vaccines addressing other deleterious enteric pathogens are needed to further reduce diarrhea-attributed mortality, morbidity, and long-term consequences. Decisions to develop, introduce, and use vaccines balance on burden, both of acute disease and associations with long-term morbidity, feasibility of developing vaccines, as well as their expected uptake and use in countries.

An expert working group convened by the World Health Organization (WHO) identified four priority enteric pathogens based on morbidity, feasibility of vaccine development, contribution to AMR, and expected vaccine uptake: *Shigella*, enterotoxigenic *Escherichia coli* (ETEC), *Campylobacter jejuni,* and norovirus.(4) *Shigella*, St-ETEC, and *Campylobacter,* are bacteria prone to antibiotic resistance that are common causes of diarrhea, growth faltering, and antibiotic use. Norovirus, a viral pathogen, is an important cause of diarrhea and antibiotic use in both high-and low-to middle-income countries also with links to linear growth faltering.

Recent systematic reviews were conducted summarizing the attribution of specific enteric pathogens to diarrhea(5) as well as the case-fatality rates of several enteric pathogens(6). We recently conducted a systematic review of the longer-term consequences of *Shigella* infection in young children, outlining evidence of this gram-negative bacterium’s impact on persistent diarrhea, linear growth faltering, and potentially catastrophic health spending(7). Here we report findings from a similar systematic review of studies evaluating the associations of ETEC, *Campylobacter*, and norovirus with growth and neurocognitive outcomes. As vaccines for these pathogens continue to move down the pipeline, this evidence summary can inform prioritization based on likelihood of averting not only diarrhea, but also growth faltering and other long-term morbidities.

## Methods

PubMed, Embase, LILACS, and SciELO were searched for articles published between January 1, 1980 to August 21, 2024 using terms outlined in **Supplementary Table 1 (Table S1)** and on Prospero (CRD42024600676). Using COVIDENCE (Veritas Health Innovation, Melbourne, Australia), titles and abstracts were reviewed by two independent reviewers using pre-specified inclusion criteria. These studies were further probed by a full text review performed by the two independent reviewers, to finalize the list of included studies. Publications had to meet all of the following criteria to be included: included children under 5 years with one or more priority pathogens (*Campylobacter*, ETEC, norovirus) identified from a fecal sample (irrespective of the presence of diarrhea); published from 1980 on; and included prospective (not cross-sectional) ascertainment of one or more outcomes of interest. Outcomes of interest included: anthropometric measurements including weight, height, length/heigh-for-age z-score [LAZ/HAZ], weight-for-age z-score [WAZ], weight-for-height z-score [WHZ], and mid-upper arm circumferences [MUAC]), stunting [LAZ/HAZ]<-2, wasting [WHZ<-2 in any age group or MUAC<12.5 in over 6 month olds]), and underweight (WAZ<-2) as well as neurodevelopmental outcomes.

The following study-level information was abstracted from included manuscripts: publication information, study design, study setting, study population, participant age-range, recruitment time frame, and pathogen detection method (bacterial culture, enzyme linked immunoassays [ELISA], and/or polymerase chain reactions [PCR]). Within a study, outcome measures and/or measures of association were abstracted for each of the following strata: time between pathogen detection and outcome ascertainment, unadjusted and adjusted (for all reported sets of confounders), by various exposure group categorization, including whether the pathogen was identified during diarrhea or sub-clinically, and by country.

We utilized a 10-point quality rating system adapted from the Strengthening the Reporting of Observational Studies in Epidemiology (STROBE) Statement(8). Points were allocated based on key metrics reported in the included article and/or in referenced parent articles, such as relating to adequate description of the study population, study design, statistical power, bias addressed through participant selection and/or statistically, and describing how missing data and losses to follow-up were accounted for (**Table S2**). A maximum score of 10 points indicated the highest quality. Scores of eight to ten were considered good quality, scores of five to seven were considered fair, and scores of less than five were considered poor quality.

## Results

We screened 3,784 non-duplicate titles and 95 full text articles, of which 30 met inclusion criteria. Several publications utilized data from the same parent study; eleven publications were from the MAL-ED study(9), six from the GEMS(10), two from the PROVIDE study(11), and two from the same surveillance study in Peru (**Table 1**, **Figure 1**). Included studies were conducted as early as 1978-1979(12) and as recently as 2020-2021. (13) When considering unique cohorts, studies were most commonly conducted in Africa (n=8) and Asia (n=5), followed by South America, with Bangladesh having the most representation among countries (n=7), followed by India (n=2), Peru (n=2), and Pakistan (n=2). Prospective outcomes were assessed, most commonly, between 2 and 24 months, with some studies including 60-month follow-up and one with up to 96 months of follow-up. Linear growth (change in LAZ/HAZ; absolute change in length; risk of stunting) was the most assessed outcome category, followed by weight gain (change in WAZ; absolute change in weight; risk of underweight), and change in WHZ/wasting. Neurodevelopmental outcomes were only assessed in two studies. The STROBE adapted quality ratings of included study scores ranged from five to ten, with 20/30 studies meeting the good quality assessment. Four(14–17) out of the five highest scores (14–18) corresponded to the manuscripts reporting from large multi-country studies on multiple pathogens (GEMS and MAL-ED)—enabling within-study comparisons across pathogens.

**Figure 1.**
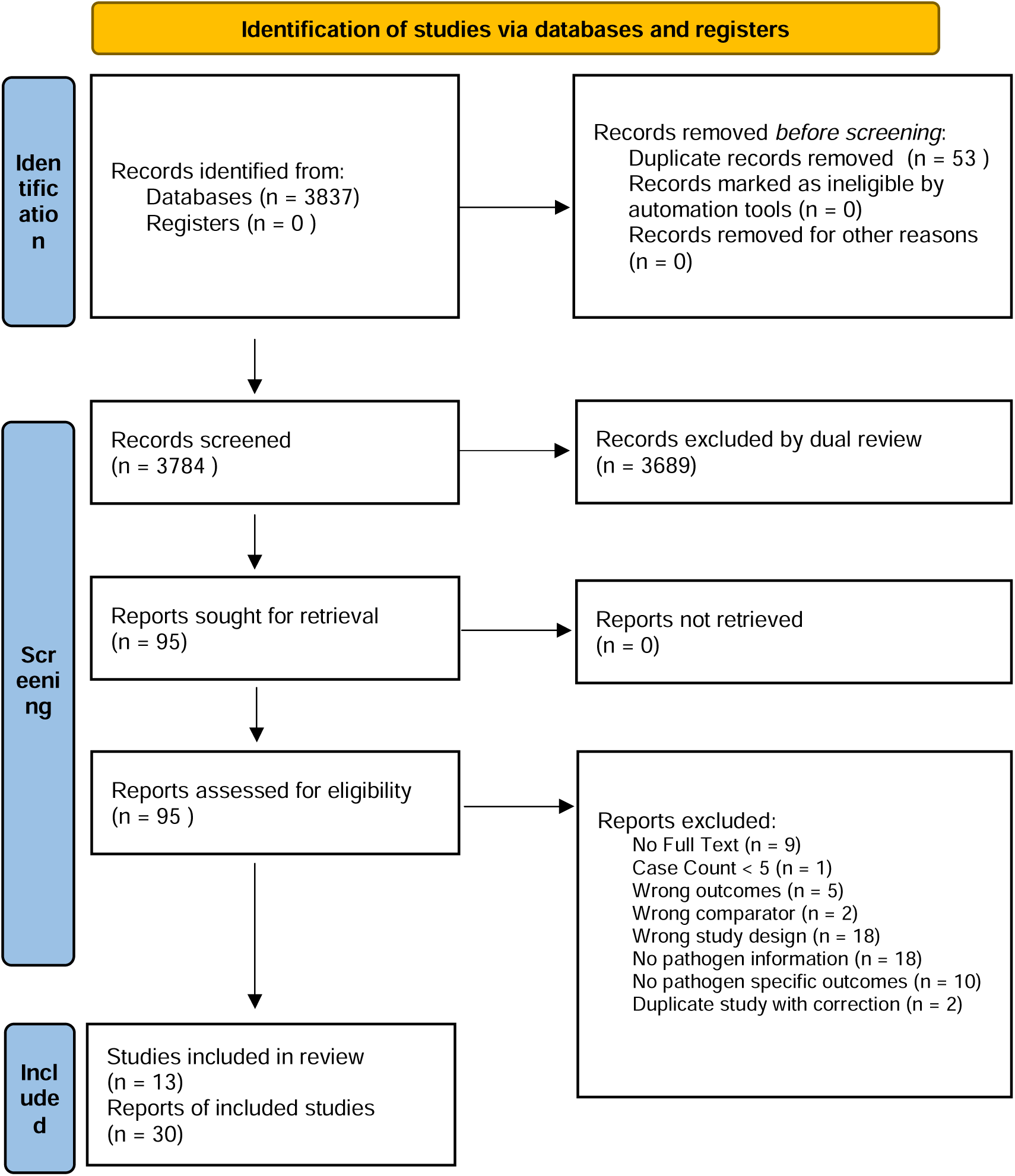
Study inclusion PRISMA Flow Diagram Source: Page MJ, et al. BMJ 2021;372:n71. doi: 10.1136/bmj.n71. This work is licensed under CC BY 4.0. To view a copy of this license, visit https://creativecommons.org/licenses/by/4.0/

**Table 1.**
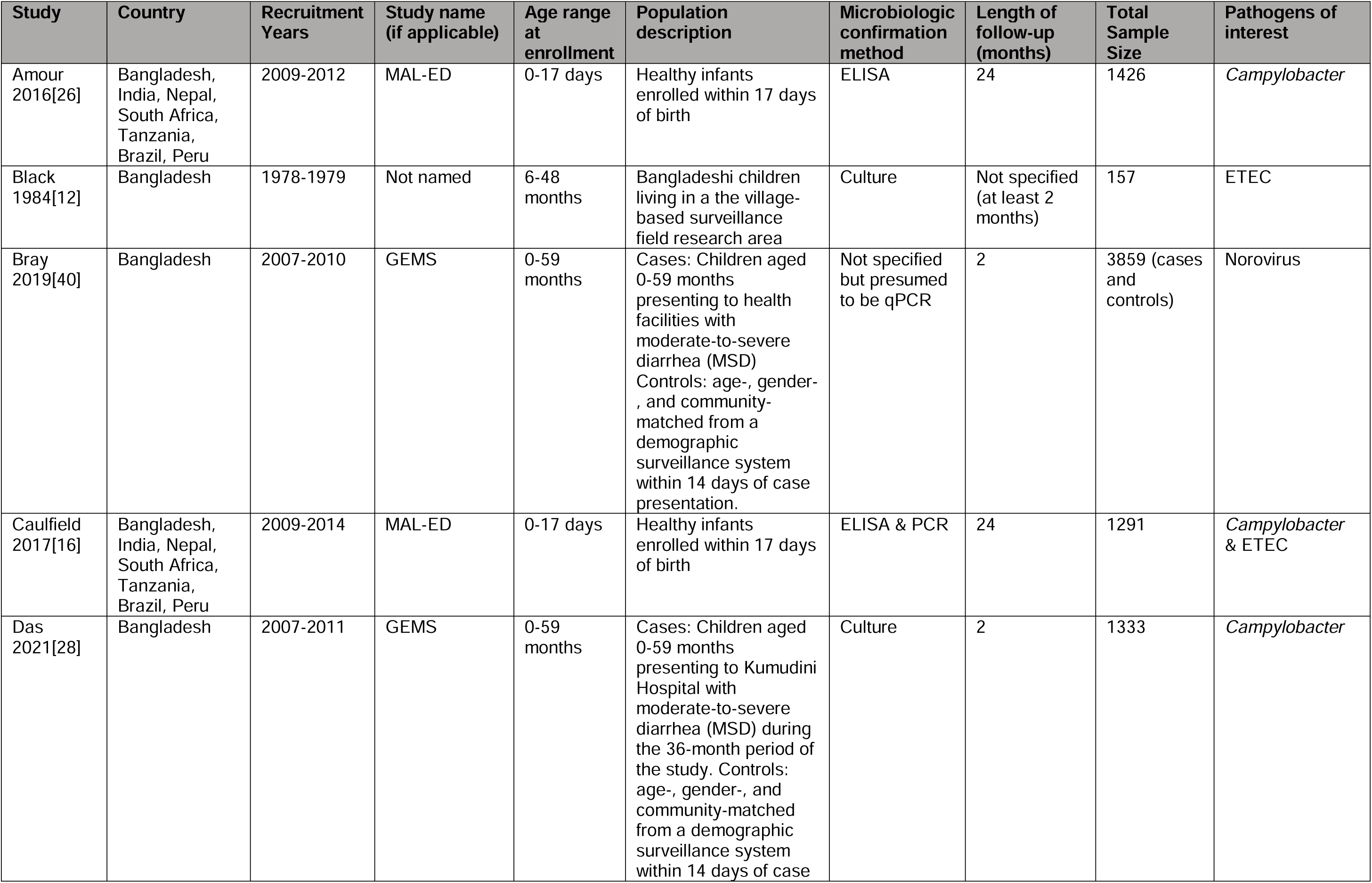
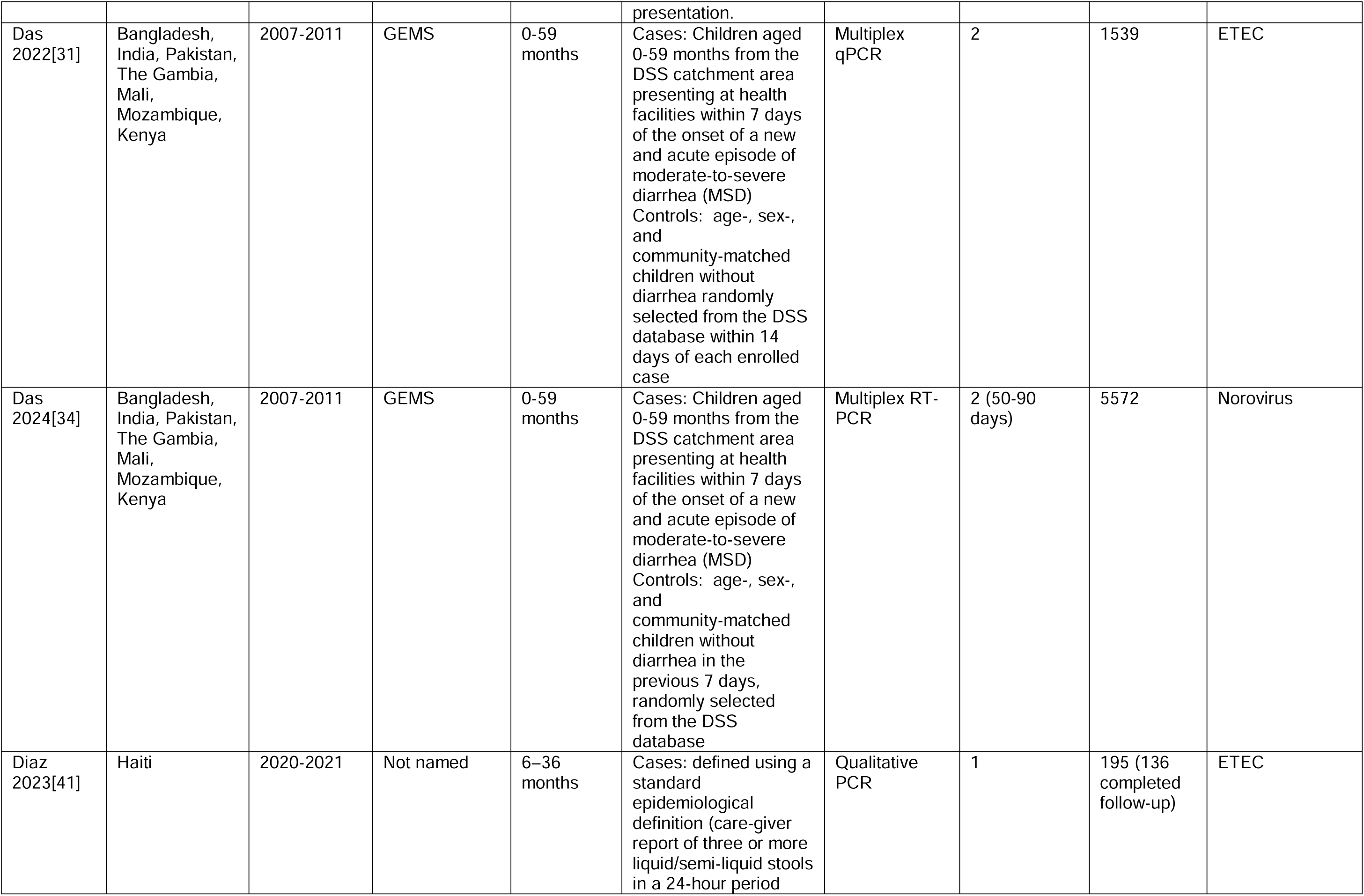
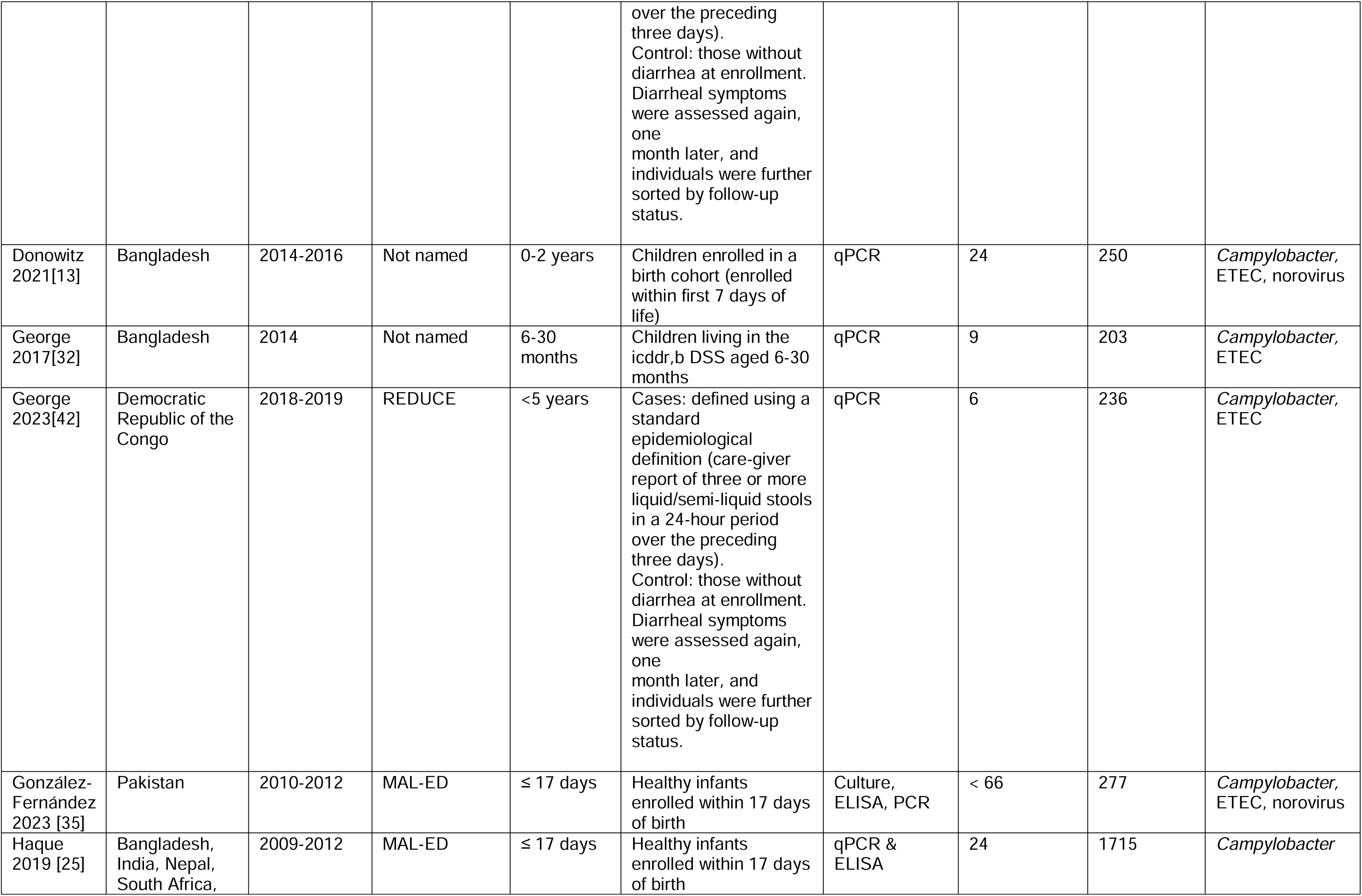
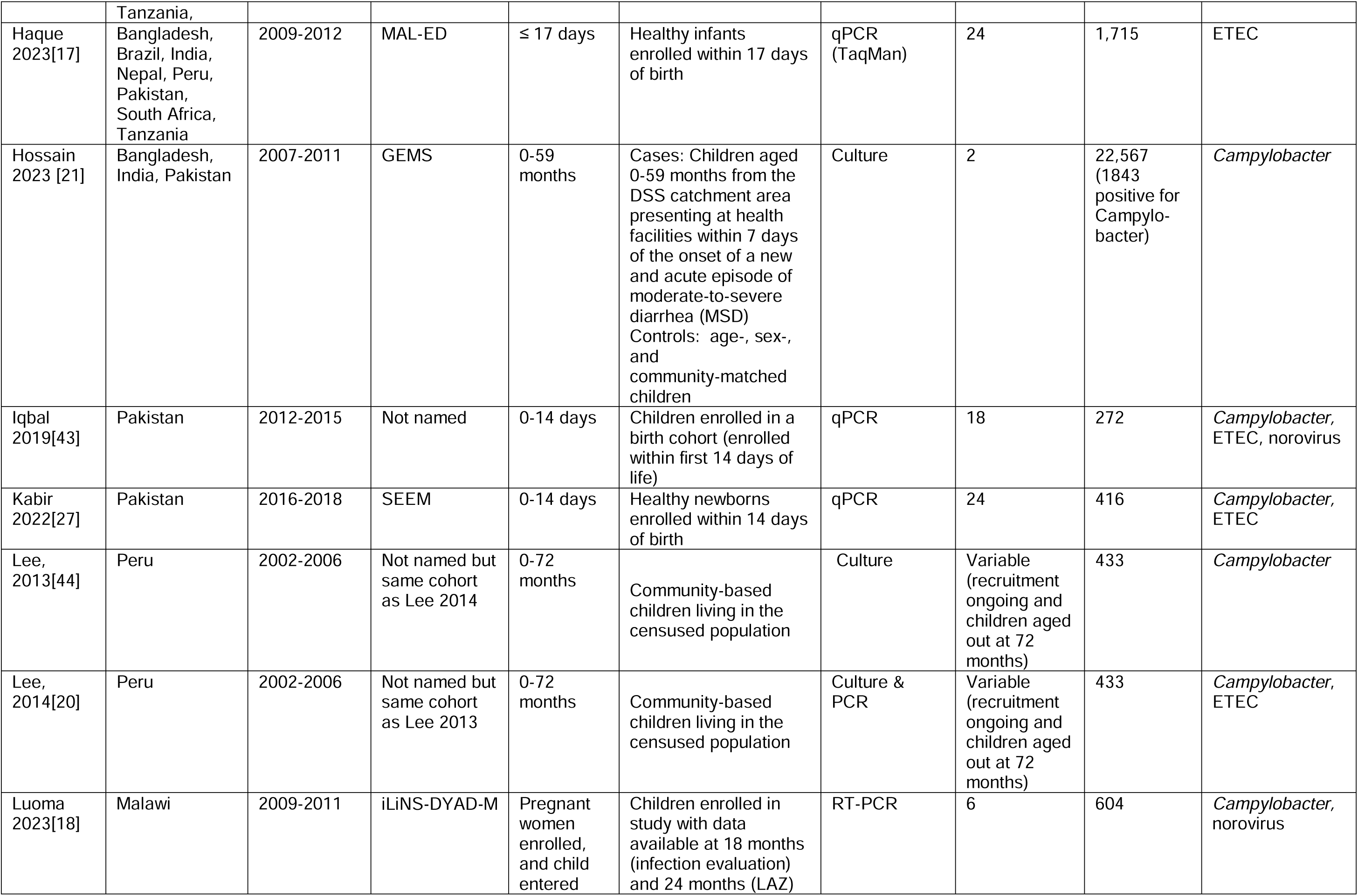
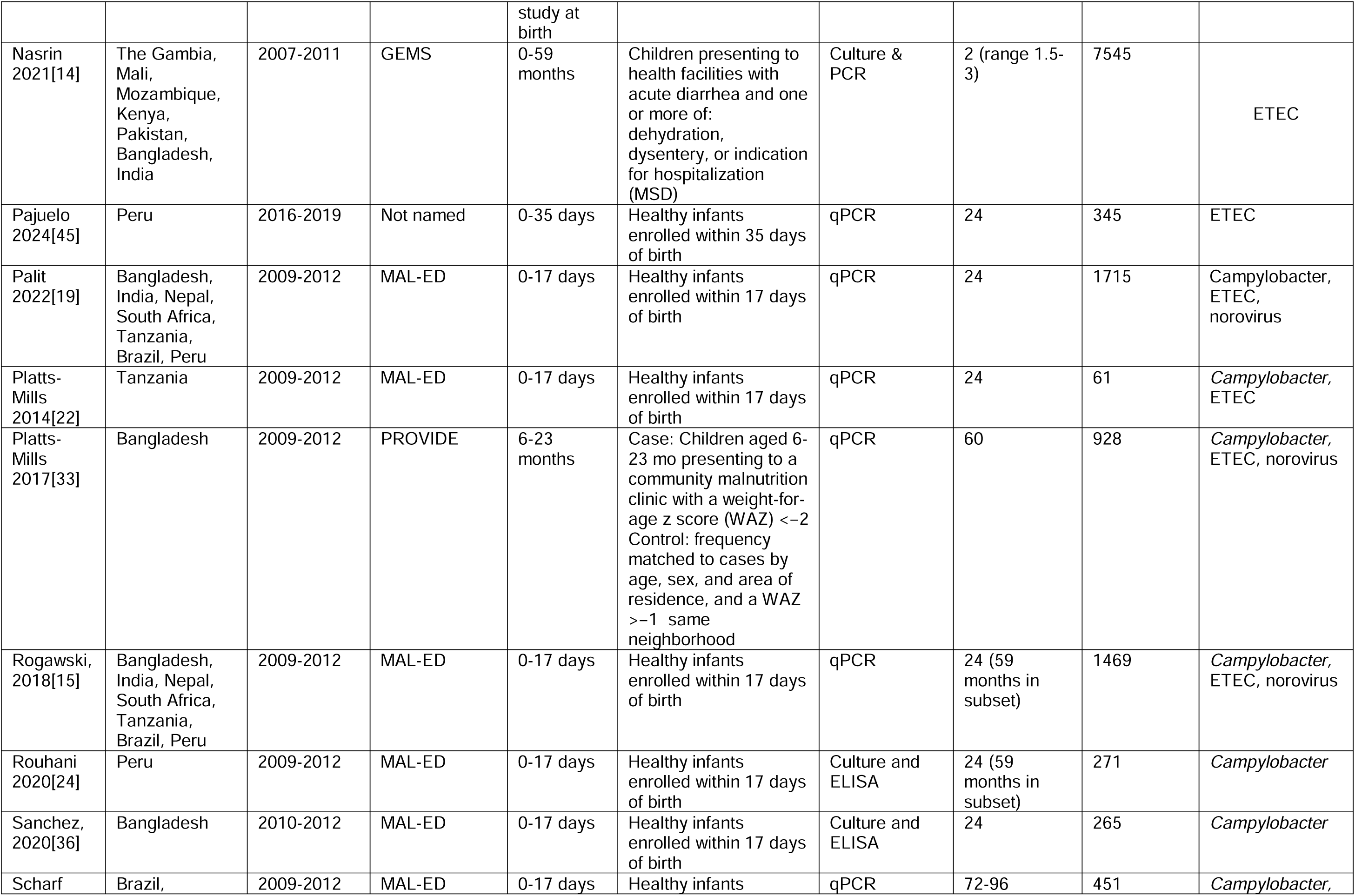
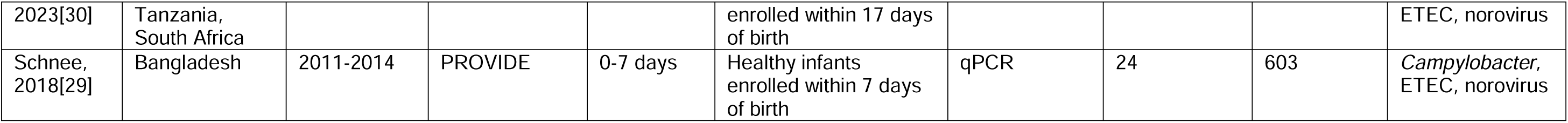
Description of included publications.

### Campylobacter

Twenty-two publications reported on consequences of *Campylobacter* (**Table 2a**), with 12 reporting weight gain outcomes, five WHZ outcomes, 20 linear growth, and two on neurodevelopmental outcomes. Four out of the 12 publications that addressed weight change as measured by absolute weight or WAZ found a statistically significant association between *Campylobacter* and weight. Each *Campylobacter* detection at routine monthly stool sample collection in the MAL-ED cohort was associated with a 0.63 (95%CI: -0.79, -0.43) lower WAZ(19) over a 24-month period. Another publication using MAL-ED data found a 0.22 lower WAZ over a 24-month period when comparing WAZ between children with *Campylobacter* detection in every monthly stool samples during predetermined time intervals to those with no *Campylobacter* infections. (16) Each incident episode of *Campylobacter* diarrhea was associated with a 55.3g (95%CI: -102.1, -8.4) lower weight two months later, but % days with *Campylobacter* from the same study population was not (-2.9g [95%CI: -7.9, 2.2).(20) Wasting was only associated with *Campylobacter* in one of the five studies that assessed this outcome, reporting an average loss of 0.16 SD in WHZ after 60 days of follow-up among children 24-59 months with asymptomatic *Campylobacter* infection(21).

**Table 2.**
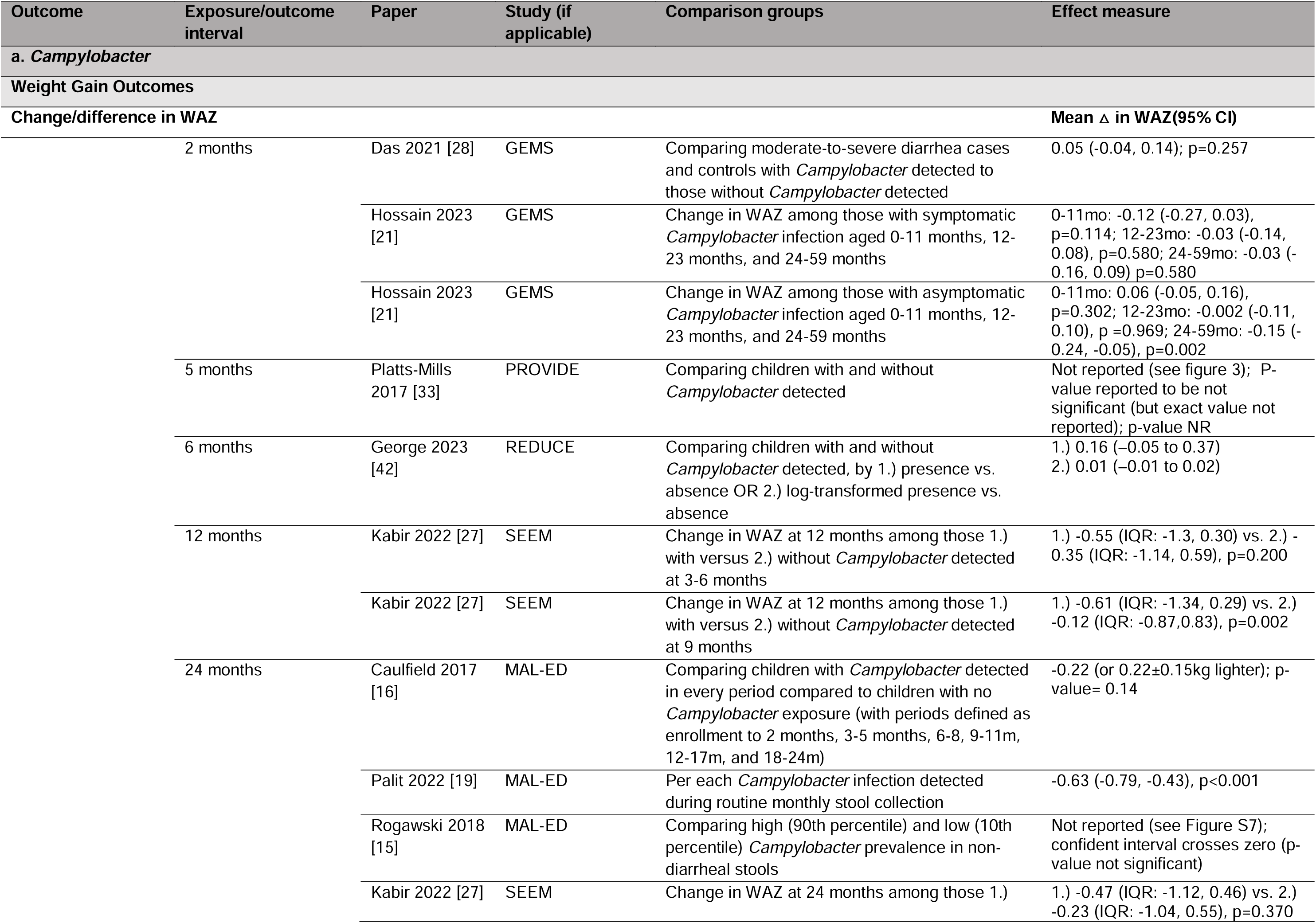
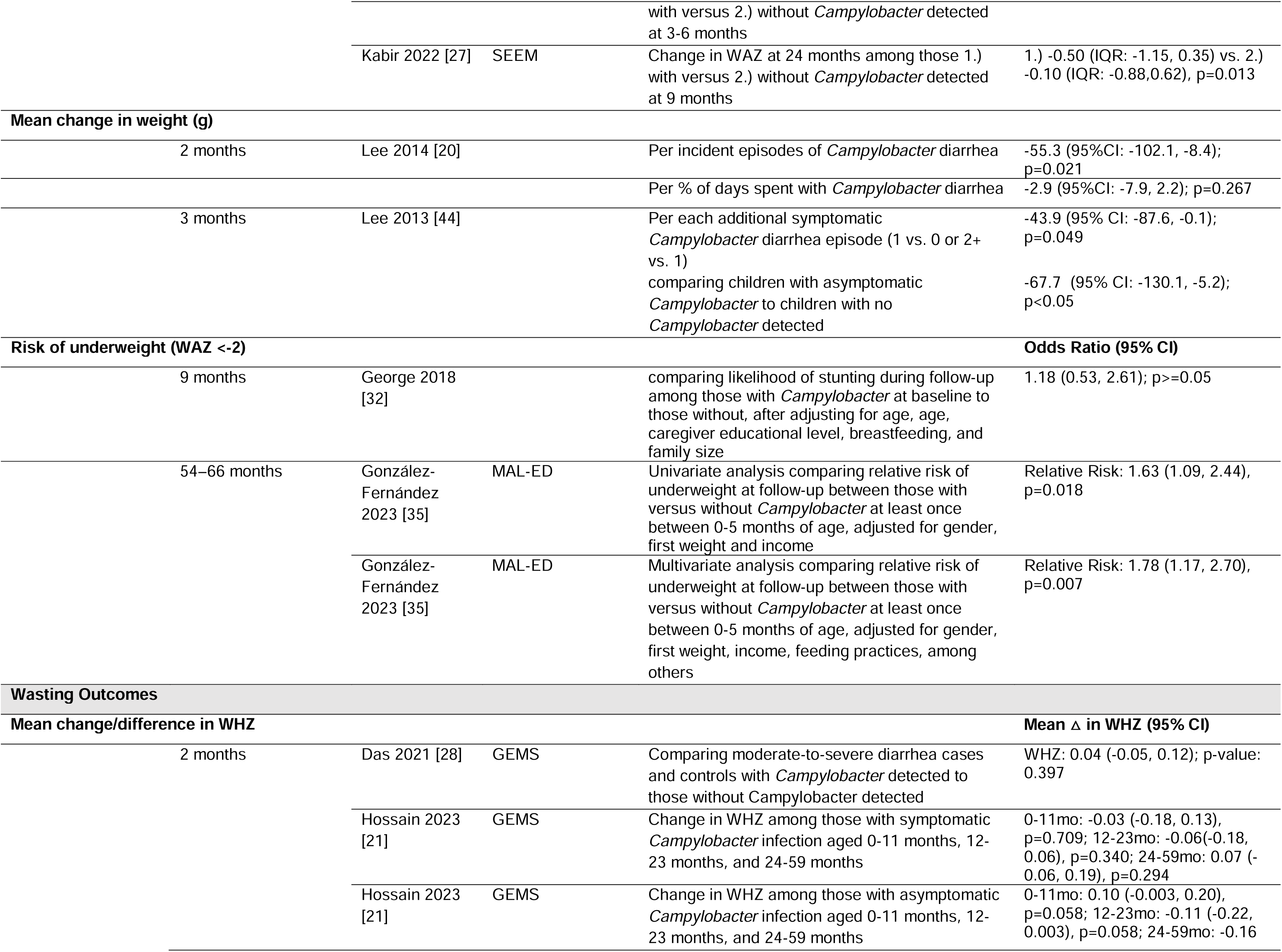
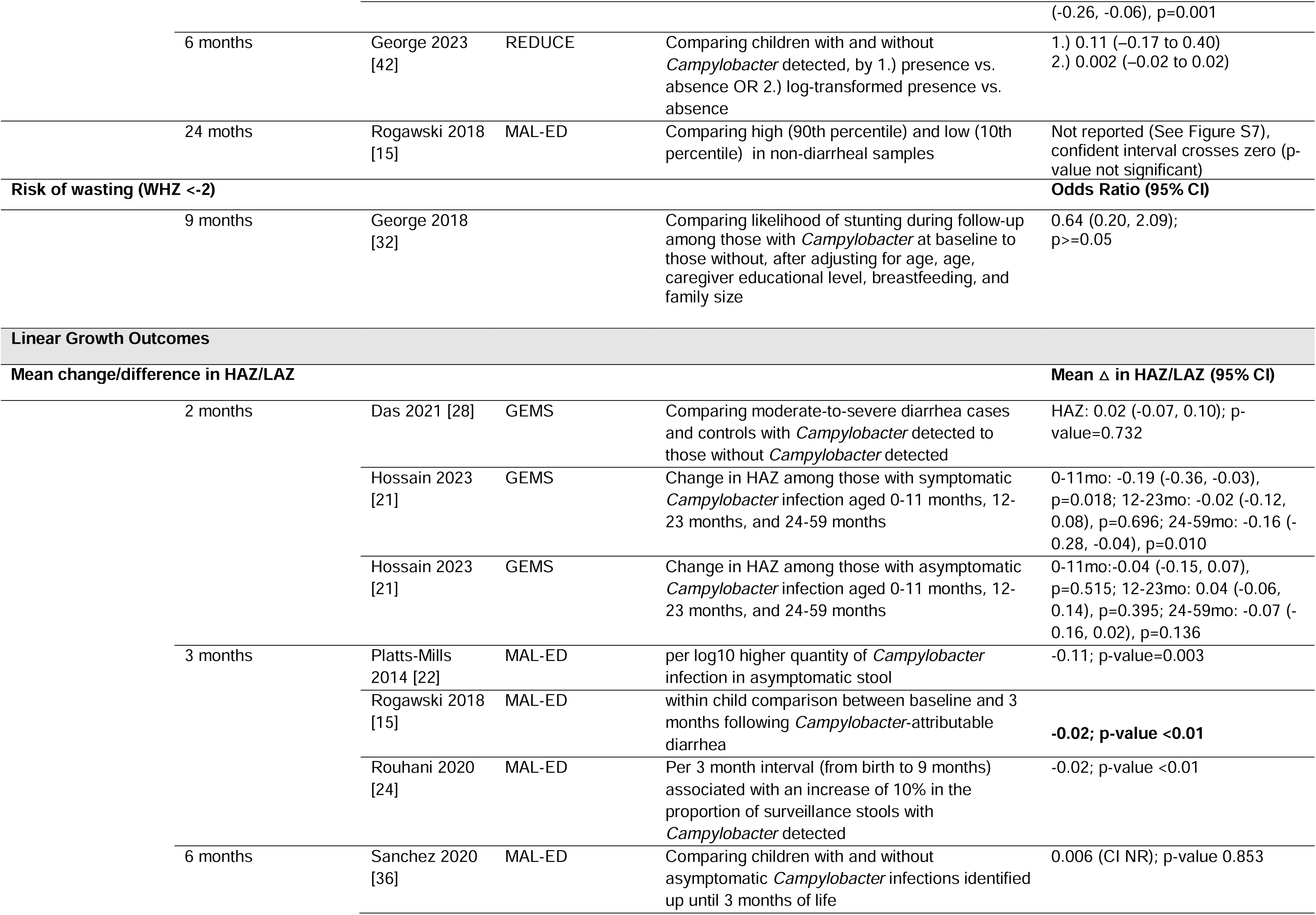
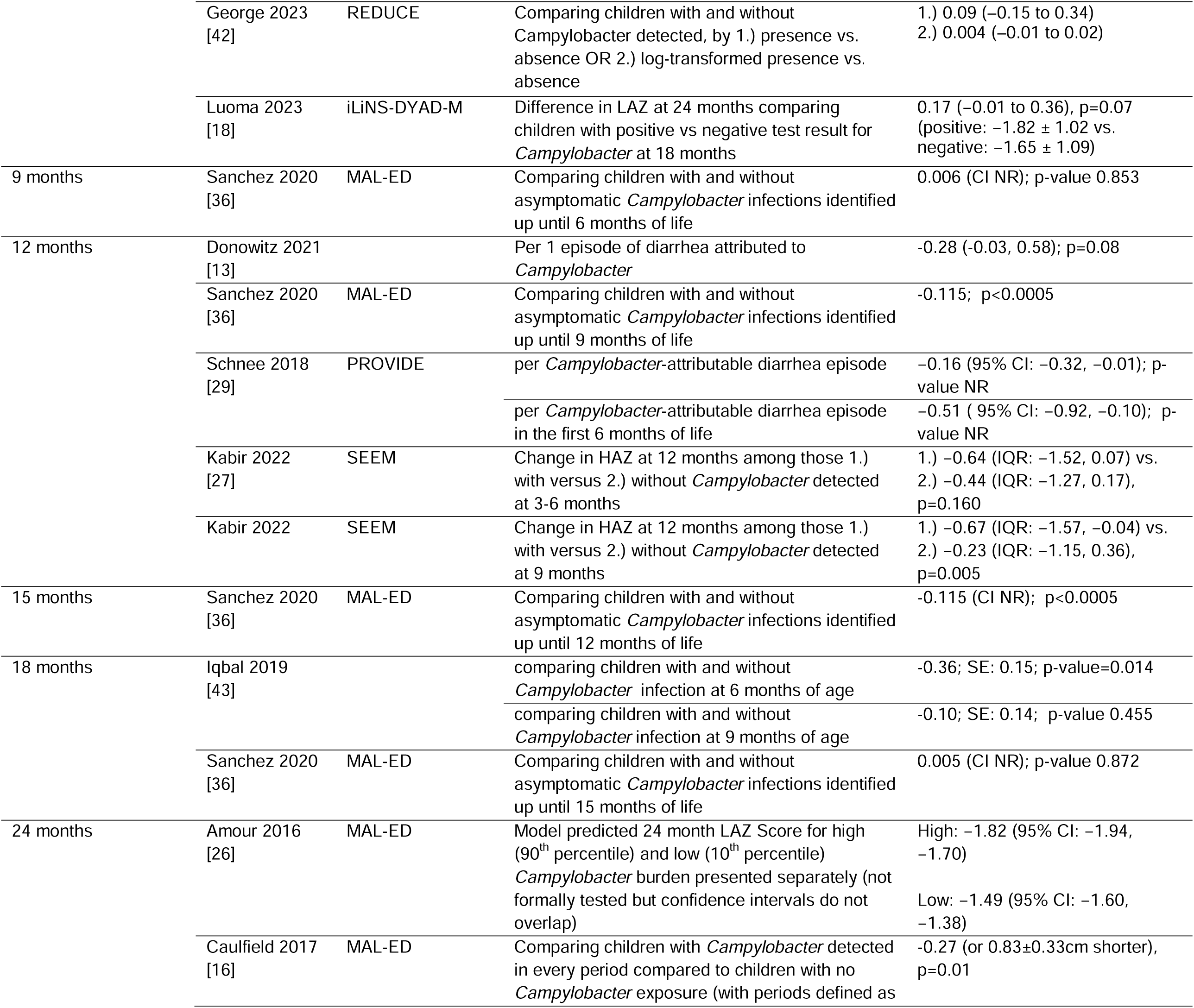
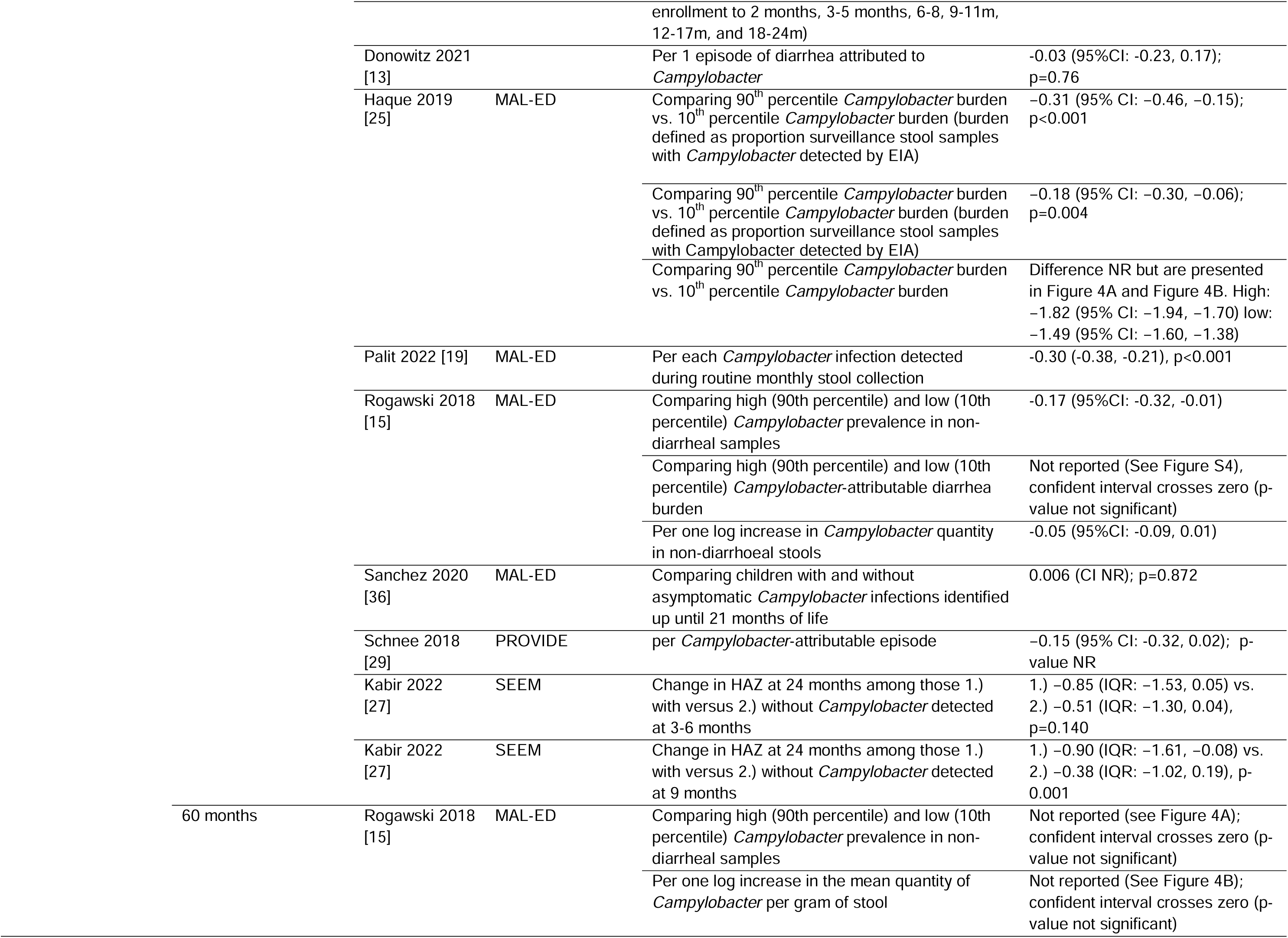
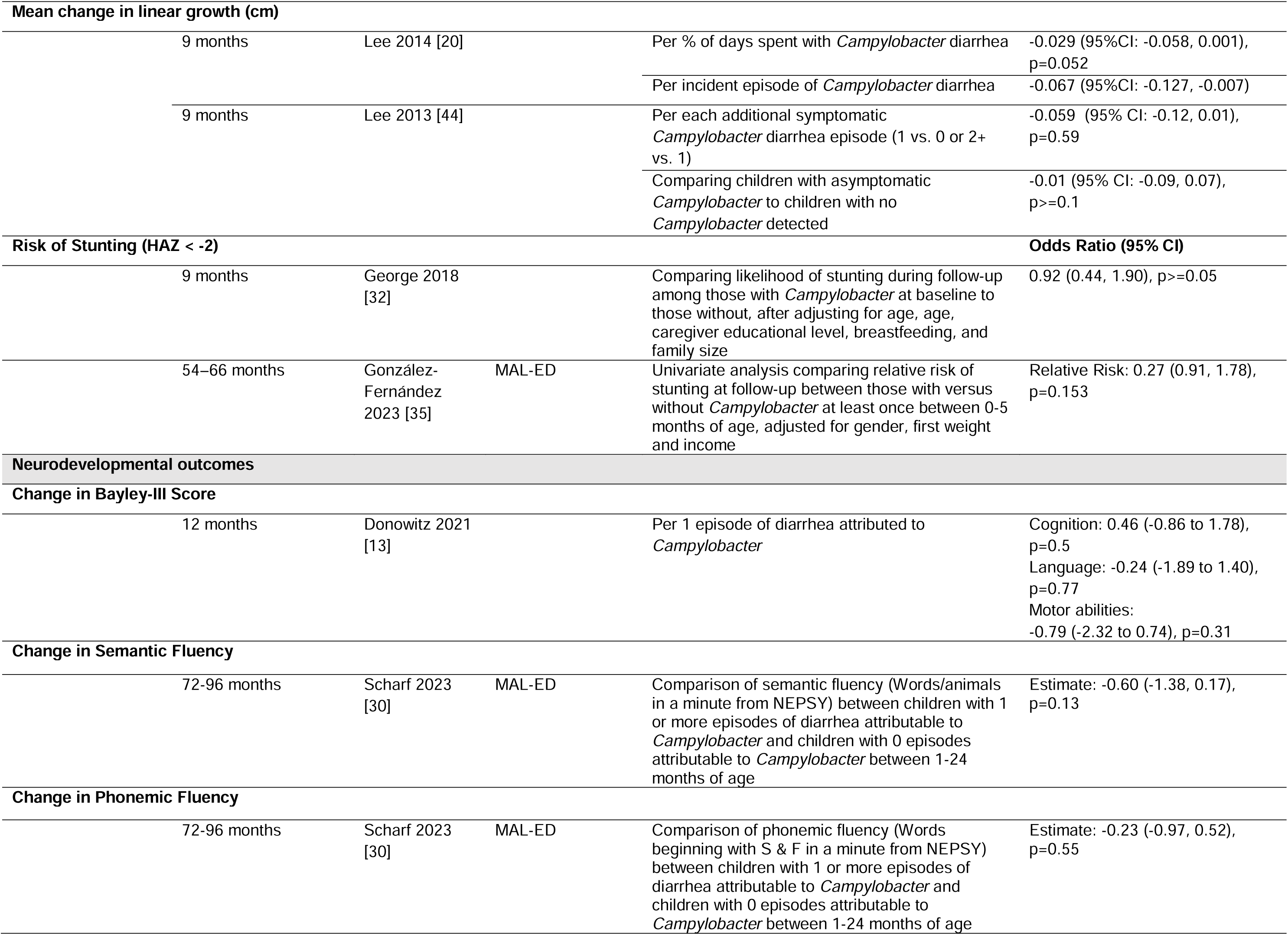
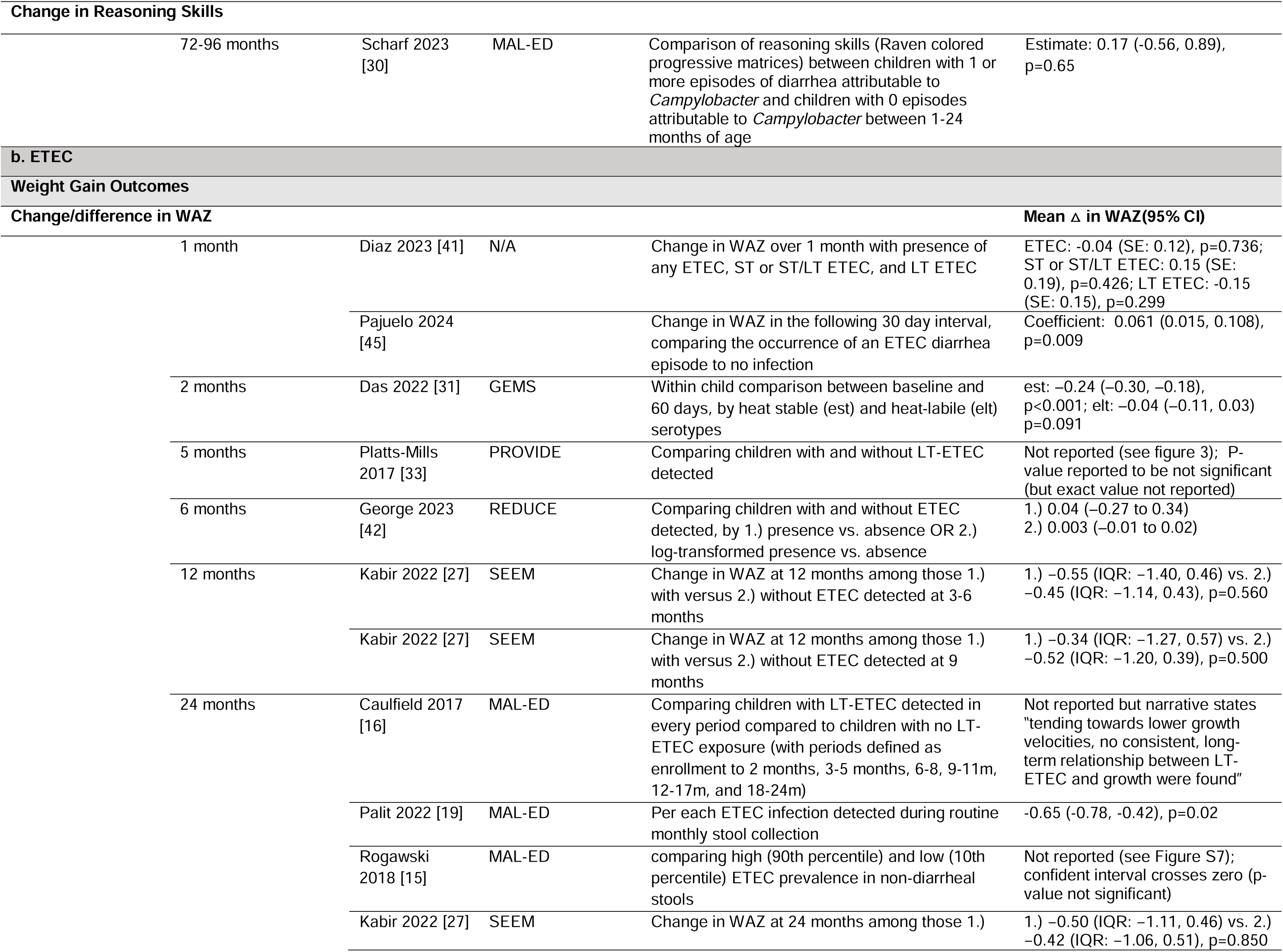
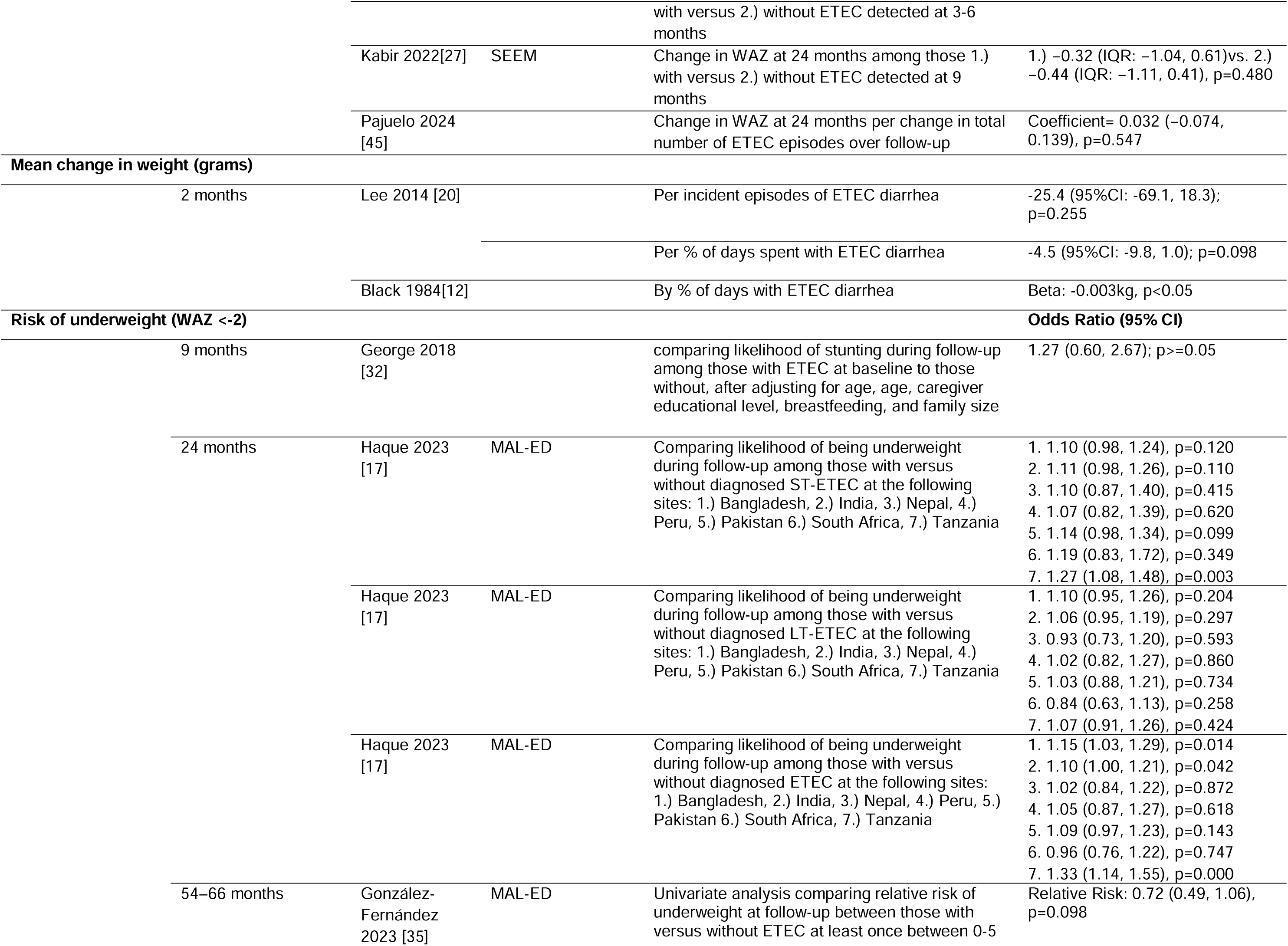
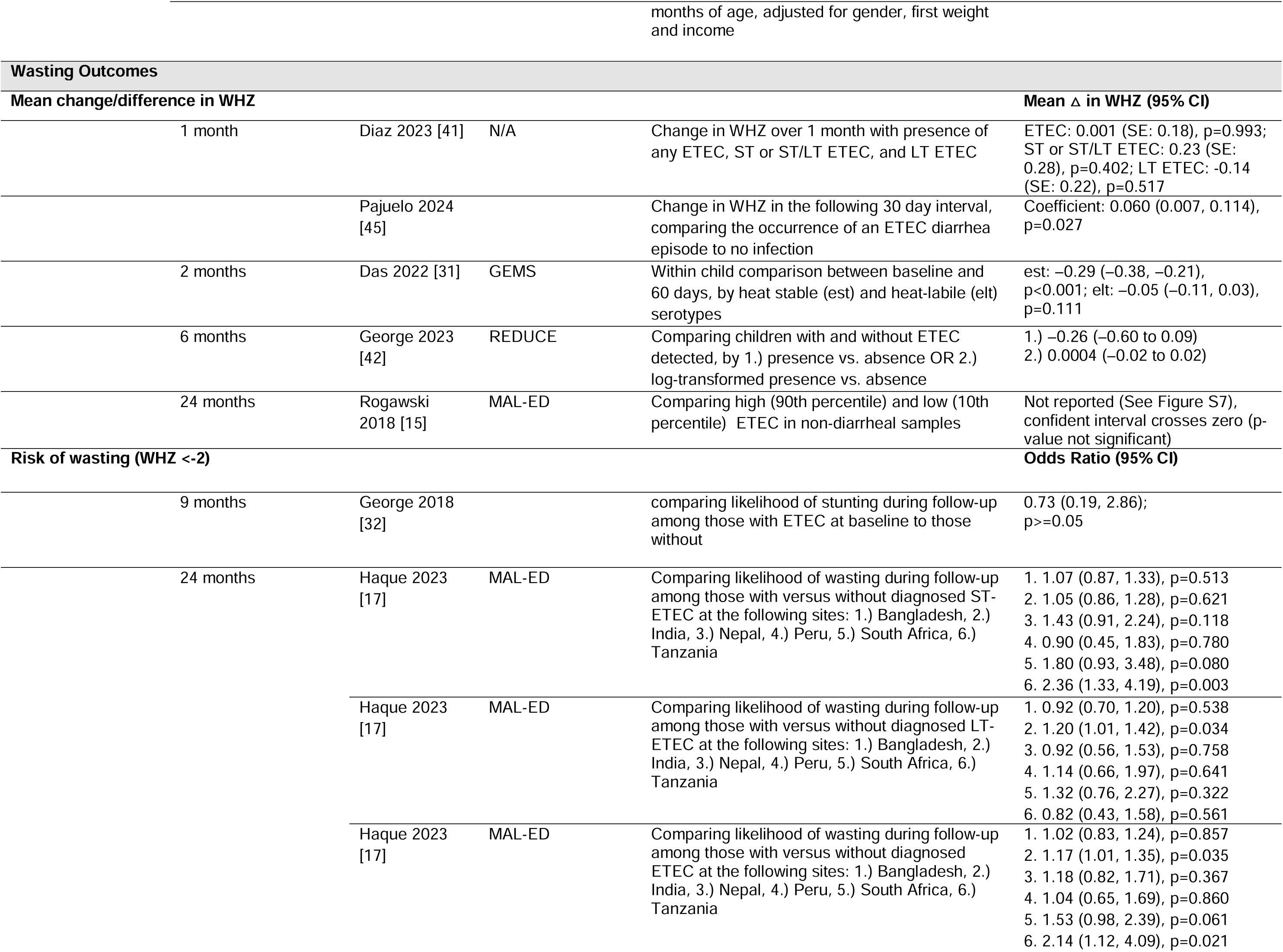
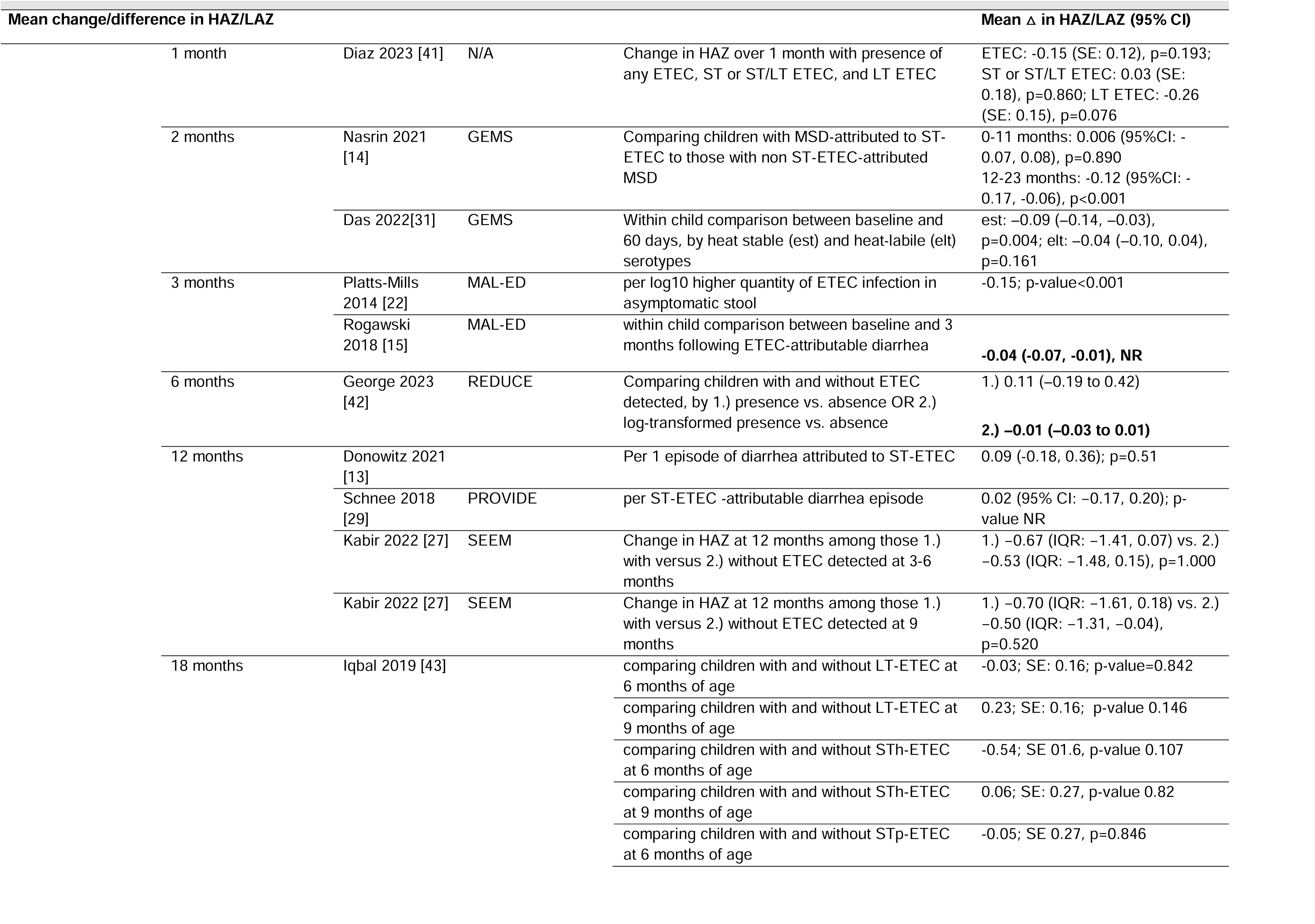
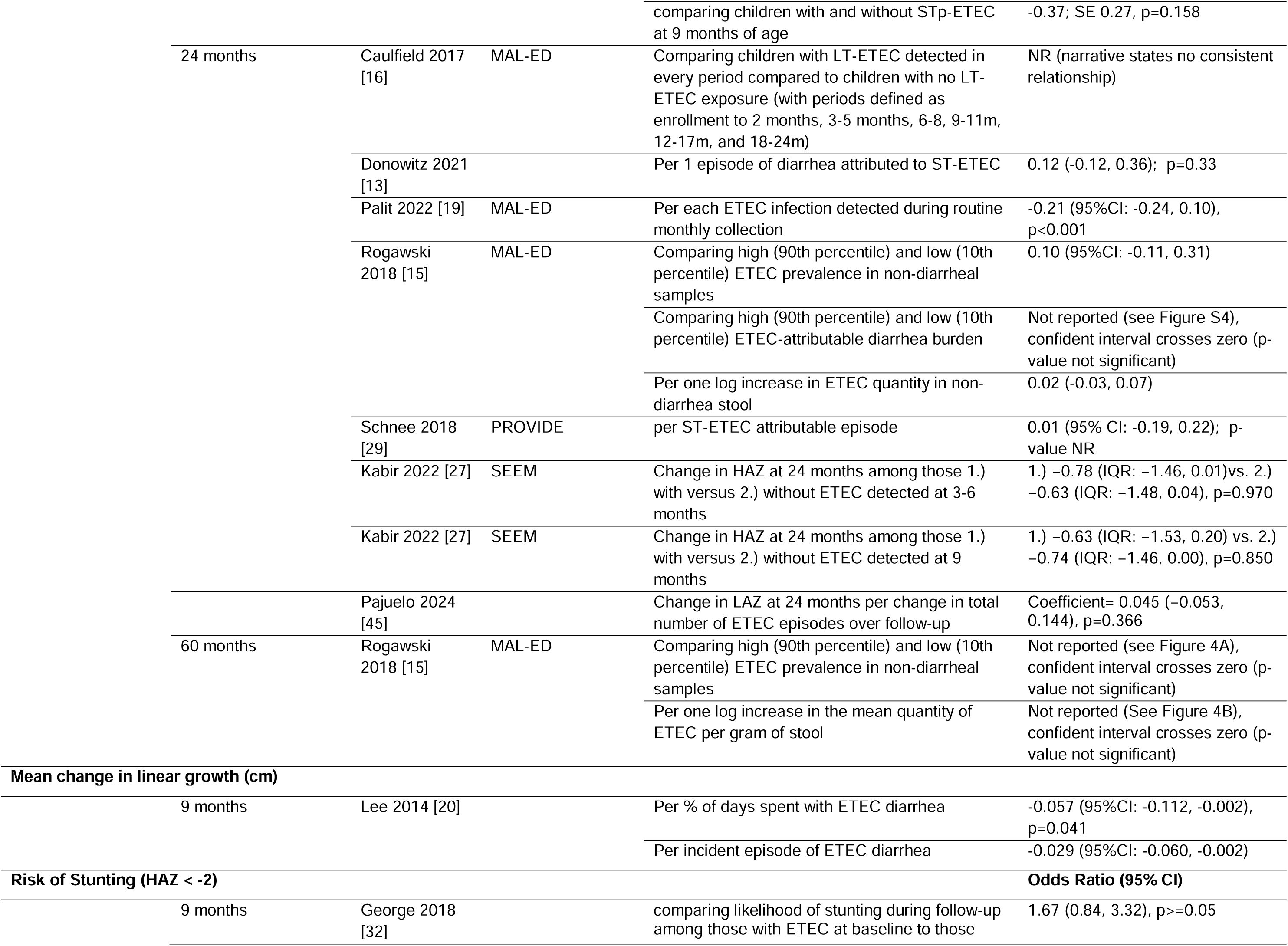
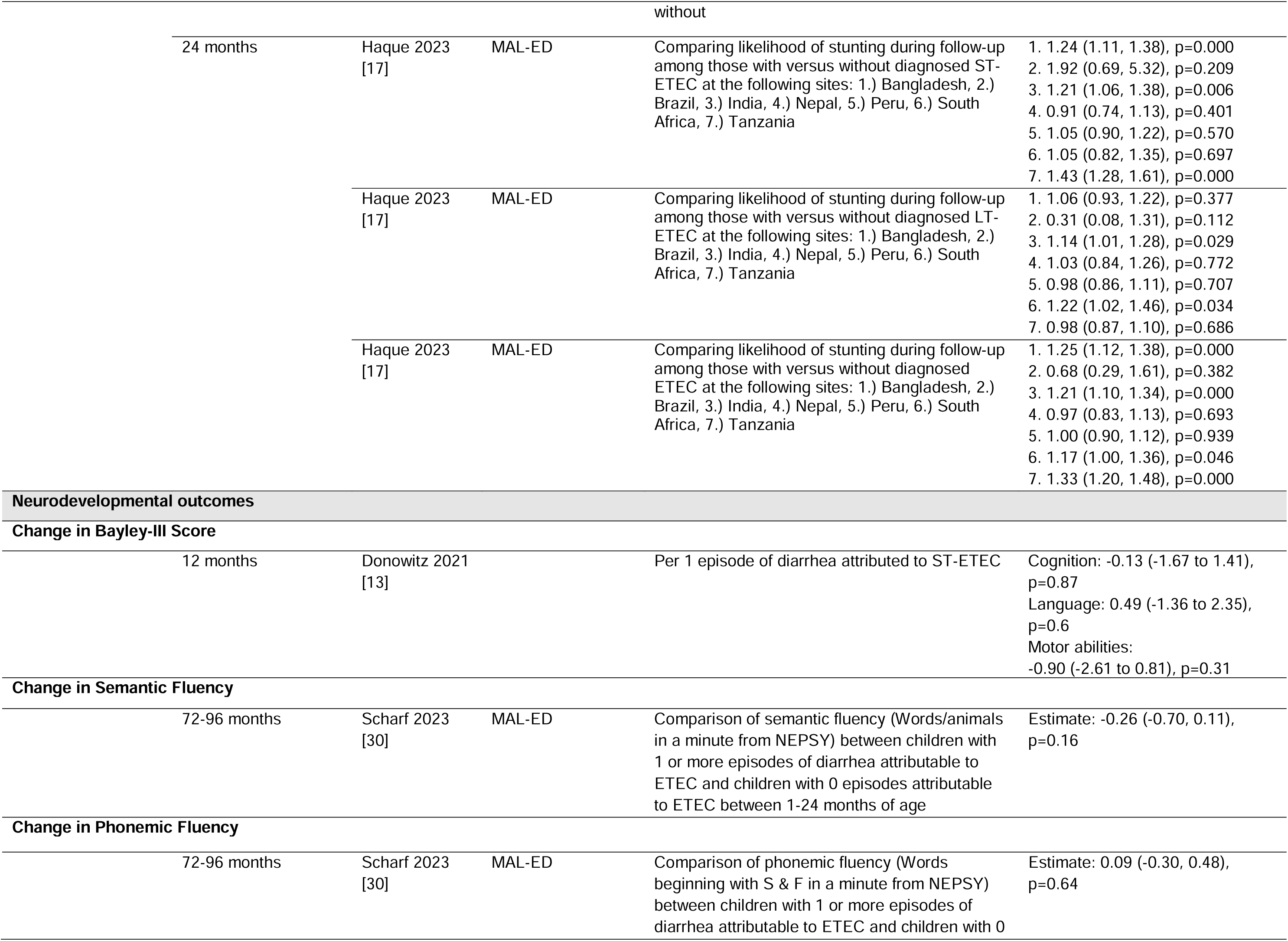
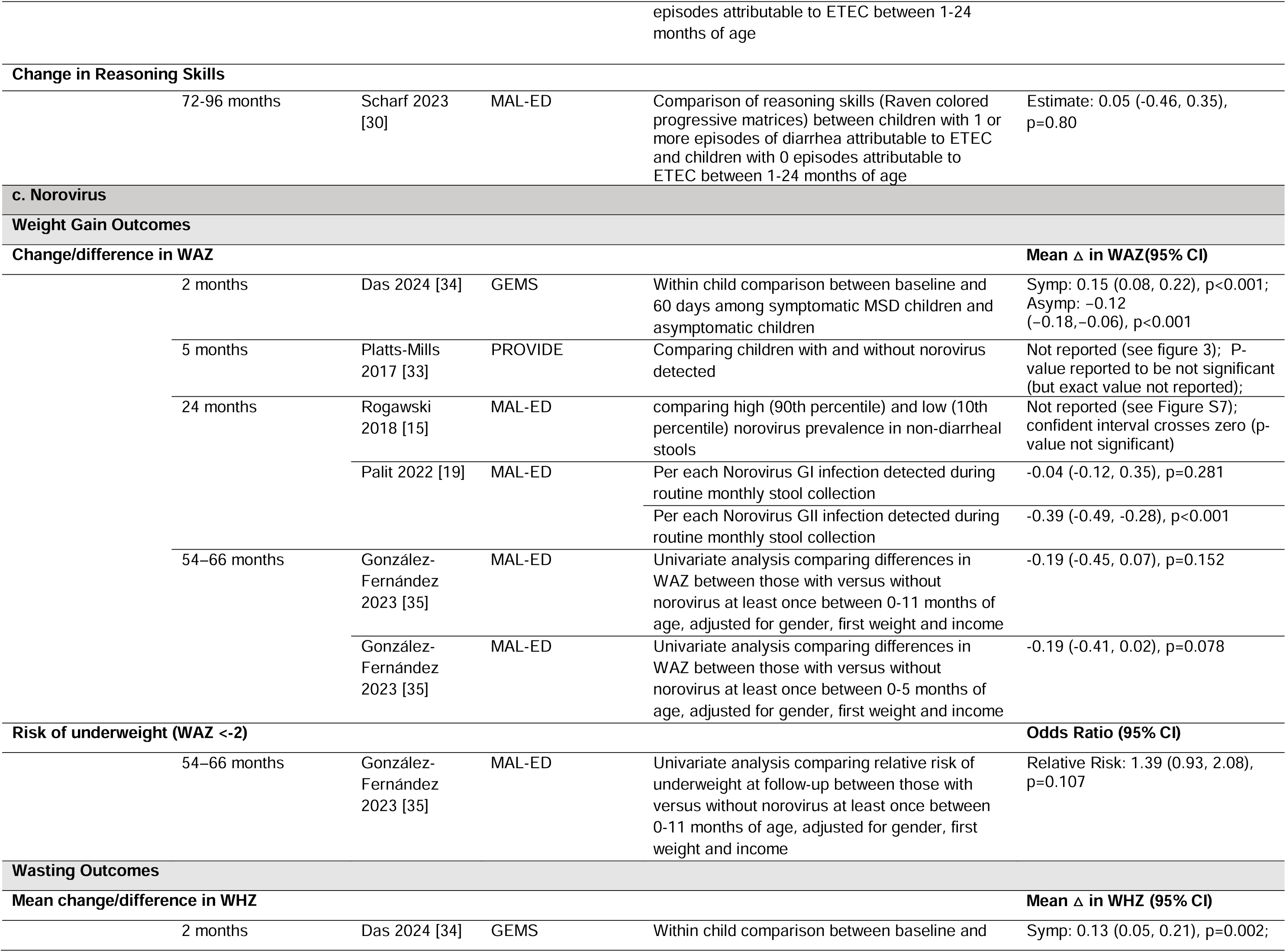
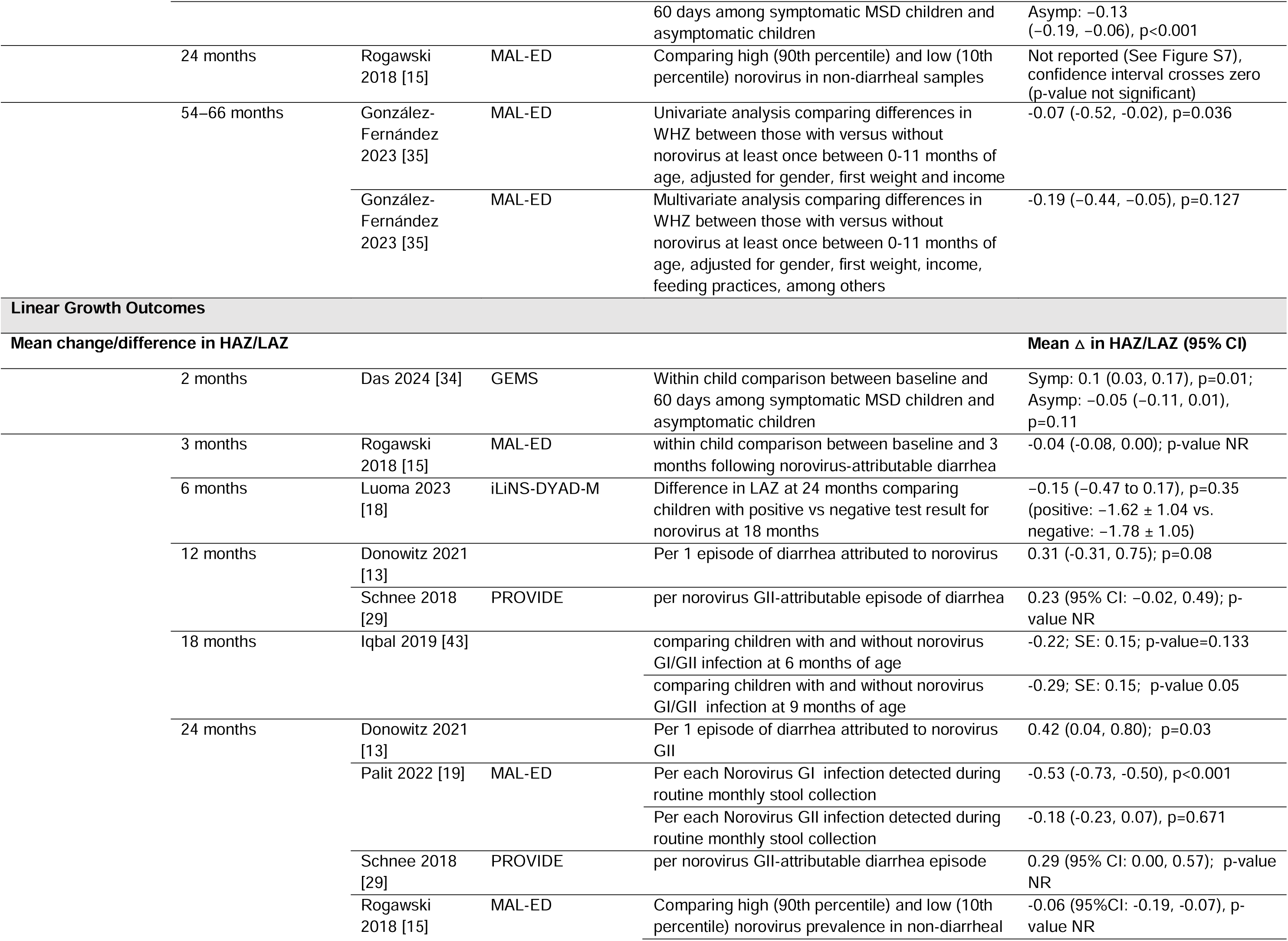
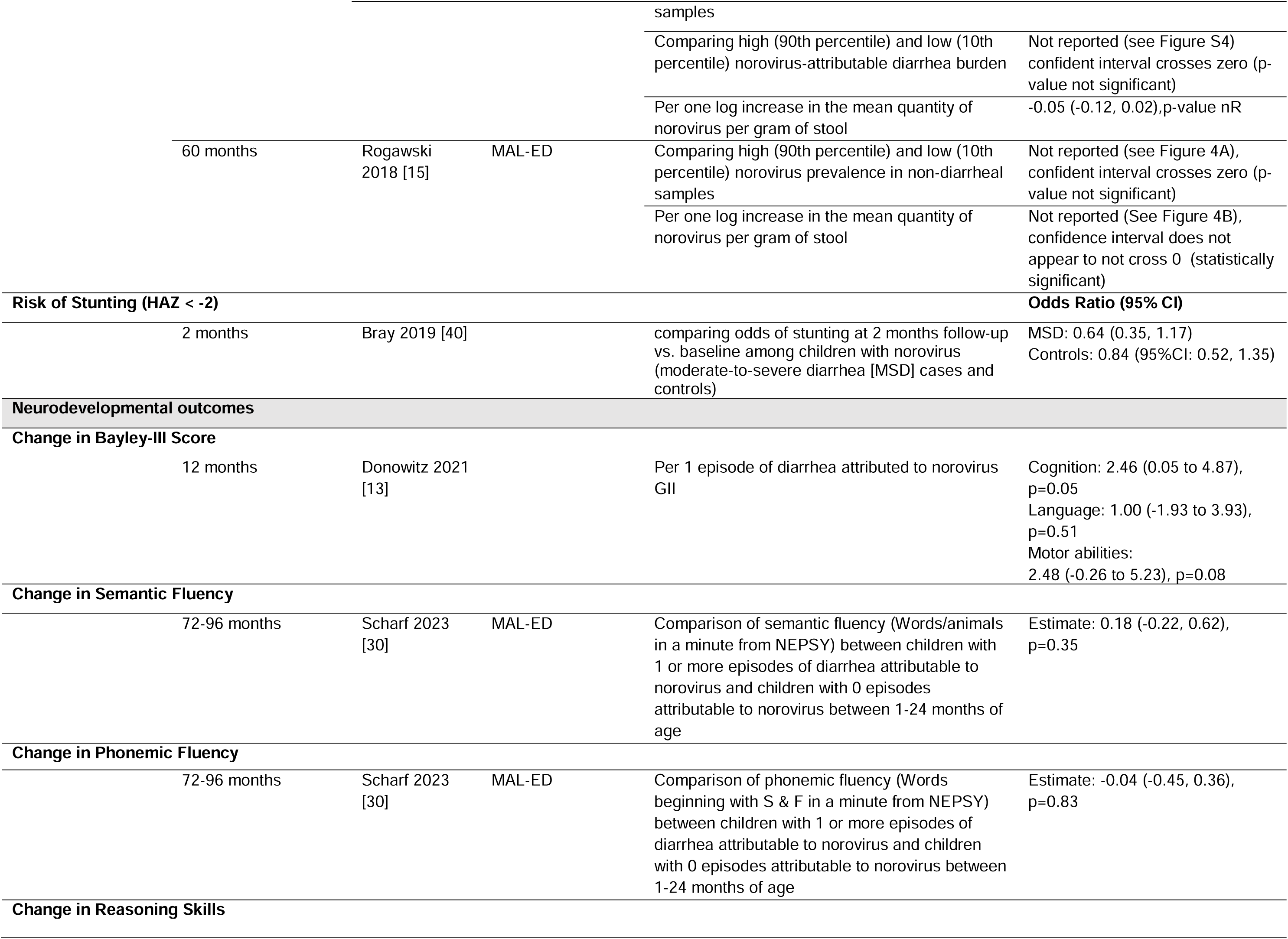
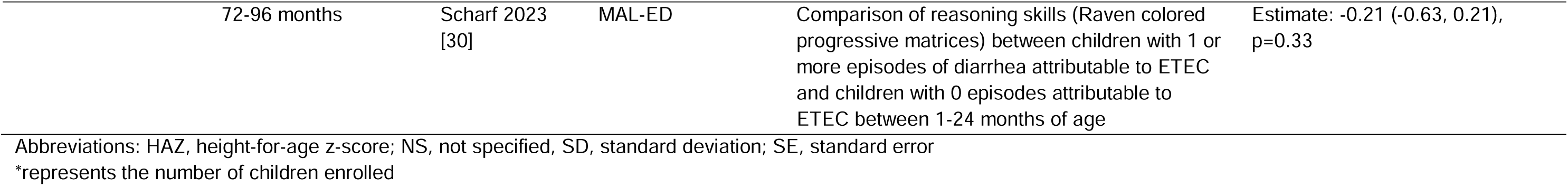
Outcomes among children with *Campylobacter,* ETEC, and norovirus, (for studies presenting multiple models for same effect [ie varying adjustments, stratification by diarrhea status, species, or country] reported least stratified adjusted model and all models presented in outcomes excel).

*Campylobacter* had some evidence of association with length change, both in the short term (3 months) (22–24) and long term (24 months) (16, 19, 23, 25–27), albeit inconsistently and effect sizes ranged from -0.90 to 0.006. Other studies found no statistically significant relationship between *Campylobacter* and linear growth. (13, 28, 29) There did not appear to be a consistent association with length measures based on whether or not the *Campylobacter* was detected with diarrhea compared to without. *Campylobacter*-attributed diarrhea was inconsistently associated with LAZ two months later(21, 28) and not at longer time intervals such as 12 (13, 29) and 24 months (13). However LAZ was modestly associated with *Campylobacter* diarrhea in MAL-ED 3 months later.(23) *Campylobacter* diarrhea was also not associated with neurodevelopmental outcomes in the two studies that evaluated this outcome. (13, 30)

### ETEC

Twenty publications reported on consequences of ETEC (**Table 2b**), with 14 reporting weight gain outcomes, seven wasting outcomes, 16 linear growth, and two on neurodevelopmental outcomes. Short term (2 months) absolute weight gain(12) was associated with ETEC-diarrhea in one Bangladeshi study but not in a study among Peruvian children(20) nor in the GEMS multicenter study(31). Each ETEC infection detected during routine monthly collection over a 24 month period in one MAL-ED publication was associated with an average 0.65 lower WAZ (95CI: -0.78, -0.42, p=0.02) (19) whereas another study in the same cohort found no consistent relationship focusing specifically on LT-ETEC.(16) Wasting did not appear to be associated with ETEC over a 9-month(32) and 24-month period(23) but site specific analyses of the MAL-ED study found an increased likelihood of wasting associated with ETEC in Tanzania(17).

ETEC was significantly associated with linear growth in three of 17 studies evaluating this association. Children aged 12-23 months (but not 0-11 month olds) with St-ETEC-attributable moderate-to-severe diarrhea (MSD) had a 0.12 greater loss in LAZ in the 60-days following their diarrhea episode than children with non-St-ETEC attributable MSD (95%CI: -0.17, -0.06), (p<0.001).(14) Cumulative asymptomatic ETEC was associated with linear growth three months after in a single site of the MAL-ED study(22) but not across all MAL-ED sites over the 24 and 60-month follow-up period (-0.04 [95%CI: -0.07, -0.01).(23) ETEC-attributable diarrhea was inconsistently associated with linear growth with a statistically significant association between ETEC-attributable diarrhea and linear growth three months after(23) in MAL-ED and in a Peruvian cohort over nine months(20) but not in two other studies looking at 12 month LAZ.(13, 29) ETEC was not associated with neurodevelopmental outcomes in the studies evaluating this outcome. (13, 30)

### Norovirus

**Table 2c** describes the eight studies evaluating norovirus’ associations with long-term outcomes (weight gain [n=5], wasting [n=3], length [n=9], and neurocognitive [n=2]). In the five studies that evaluated norovirus’ association with weight gain, one from the MAL-ED cohort demonstrated evidence of a statistically significant association between norovirus GII infection detected during routine monthly stool collection in 24 month change in WAZ (-0.39 [95%CI: - 0.49, -0.28], p<0.001) (19) The other MAL-ED cohort publication did not find an association between high (90^th^ percentile) prevalence of norovirus in non-diarrheal stools and WAZ (23) nor did the analysis from the PROVIDE study comparing change in WAZ five months after norovirus (or no norovirus) detection.(33) A GEMS secondary analysis found discrepant associations between norovirus detection and WAZ 60-days later-norovirus MSD cases gained WAZ and asymptomatic controls with norovirus infection lost WAZ (34). The two studies from MAL-ED found no evidence of association between norovirus and ponderal growth (23, 35).

Of the 18 associations reported on the relationship of norovirus on LAZ/HAZ/stunting from eight unique publications, four were statistically significant and effect sizes for associations with change in LAZ/HAZ ranged from -0.53 to 0.31. For example, mean change in LAZ between baseline and three months following norovirus-attributed diarrhea was associated with a 0.04 lower delta LAZ (95% CI: -0.08, 0.00). (23) Another analysis from MAL-ED found an association between each norovirus GI infection detected during routine monthly stool over a 24 month period and change in mean change LAZ (-0.53 [95%CI: -0.73, -0.50]) but not norovirus GII infection (-0.18 [95%CI: -0.23, 0.07]) (19). Each episode of Norovirus GII-attributed diarrhea was associated with a higher delta LAZ at 24 months (0.04 [95%CI: 0.04, 0.80). (13) Of the three pathogens evaluated in the review, norovirus (specifically norovirus GII) was the only pathogen significantly associated with a neurodevelopmental outcome, albeit in a positive direction: each episode of norovirus GII diarrhea was associated with a 2.46 higher Cognitive Bayley score (95%CI: 0.05, 4.87) (13) but had no association with semantic or phonemic fluency or reasoning skills(30).

## Discussion

The consequences of enteric pathogens beyond diarrhea are an important consideration for vaccine development prioritization. In this systematic review of observational studies following children with and without *Campylobacter*, ETEC, and/or norovirus infections, we found modest evidence of linear growth detriments associated with all three pathogens, modest evidence of *Campylobacter* impacting weight, and no evidence of detrimental impacts of these pathogens on wasting or neurodevelopment, albeit these two outcomes were rarely reported. Differences in outcome definitions, comparison groups, and timeframes prohibited meta-analysis and emphasize the need for more standardized reporting of anthropometric and neurocognitive outcomes following enteric pathogen exposure. Because these outcomes have multiple causes and occur over long-time frames, these associations are particularly prone to confounding, reverse causality, and selection bias. Ultimately, highly efficacious randomized controlled trials of interventions targeting specific enteric pathogen infection and disease are needed to establish the magnitude and relative importance of long-term consequences from enteric pathogens.

Studies conducted in the same setting frequently had discrepant results. For example, three studies evaluated linear growth over 12 months associated with *Campylobacter* in Bangladesh(13, 29, 36). Some found an association with the pathogen and outcome in the presence(29) and absence of diarrhea(36), while others did not.(13) We also found several publications utilizing the same underlying dataset (such as from MAL-ED). For example, Palit (2022)(19), Caulfield (2017)(16), and Rogawski(23) (2018) all reported on changes in LAZ 24 months after enrollment associated with ETEC from all country sites in the MAL-ED study, with change in HAZ/LAZ estimates ranging from 0.10 to -0.21 and no statistically significant association(16, 23) to a statistically significant association.(19) While these publications captured different pathogen exposure classifications (per each ETEC infection during monthly stool collection; comparing high to low prevalence in monthly stool; children with and without asymptomatic ETEC per 3 month period) and two studies used molecular methods while one used culture-based methods, the large variation in results from the same cohort was surprising. As definitive answers around relative contribution of specific pathogens to long-term outcomes is necessary to prioritize resources, consensus for the most relevant/ interpretable exposure classification and outcomes measures/timepoints would enable more efficient and interpretable evidence synthesis.

Exposure classification (per % day spent with pathogen-specific diarrhea, per incident episode of pathogen-specific diarrhea, with/without pathogen of interest at a single, prior time point, high [90^th^ percentile] to low [10^th^ percentile] pathogen-attributable diarrhea burden, per one log increase in cumulative pathogen quantity, per each pathogen detection over several time points) was not the only source of heterogeneity between, and within, studies. Differences in diagnostics (ELISA, culture, or the most sensitive, qPCR) for pathogen detection; detection of pathogens in diarrhea vs. asymptomatic fecal samples, differences in timeframes (ranging from 2 to 60 months), differences in outcome/dependent variable framing (for example, change in LAZ, single time point LAZ, change in growth (cm), odds of stunting), and differences in confounder adjustment make cross-study comparisons challenging. Individual patient data meta-analyses, an increasingly appreciated gold-standard in evidence synthesis, could overcome some challenges with sources of heterogeneity, such as comparison groups and confounders, as long as comparable information is available between studies.

Overcoming measured and unmeasured confounders remains a challenge in observational studies of long-term processes. Randomizing children to highly efficacious pathogen specific vaccines or treatment and evaluating outcomes (vaccine or treatment probe studies) will enable more robust causal inference in establishing enteric pathogen-attributable long-term morbidities by preventing confounding. The Antibiotics for Children with severe Diarrhea (ABCD) trial, a 7-country double-blind placebo controlled trial testing the efficacy of azithromycin among children with watery diarrhea and other severity indicators, found 3-days of azithromycin treatment to reduce the loss in LAZ 90-days following ST-ETEC attributed diarrhea by 0.08 (95%CI: 0.01, 0.14) compared to placebo among 889 children.(37) Too few children had *Campylobacter* identified in this trial to conduct this same probe. Whether the short-term improvements in LAZ associated with azithromycin are sustained following ST-ETEC infection, and whether such improvements translate to important developmental milestones, like neurocognitive development, remains unknown. Antibiotic probe studies are a useful tool for interrogating bacterial pathogen consequences, but to the best of our knowledge, no such treatment probes exist for viral infections such as norovirus.

While pathogen-specific vaccine trials offer the best way to obtain unconfounded estimates of long-term pathogen-specific effects they will not be without challenge. Vaccines take decades to be developed. The most advanced vaccine candidate for ETEC is in phase 2 clinical development, phase 3 for norovirus, and to the best of our knowledge, there are no candidates for *Campylobacter* at this time. Phase 3 vaccine trials are cost-prohibitive, particularly those that include lengthy follow-up to accrue long-term outcomes. Vaccines may not cover all serotypes of a given pathogen, may only prevent disease and not infection, and/or may be sub-optimally effective. Furthermore, population-level effect sizes of interventions, such as vaccines, on longer-term outcomes are expected to be small because only a minority of children in the trial will be infected with the targeted pathogen and even fewer will develop disease from said pathogen. Therefore, the overall vaccine effect will be heavily diluted by children who were not at risk for vaccine preventable, pathogen-attributable, long-term outcomes of interest, such as growth or neurodevelopmental faltering. Vaccine trials that have been powered to a primary disease endpoint are likely to be dramatically underpowered for growth and other long-term outcomes.

Longitudinal studies of early childhood diarrhea and longer-term cognitive outcomes have not shown consistent evidence of an association.(38) However, early growth faltering and biomarkers of enteric and systemic inflammation, processes likely causally linked to enteric pathogens, do appear to have stronger ties with cognitive outcomes, albeit variably by site.(30) Because the cost of following a cohort of individually randomized participants for several years is prohibitive, post-vaccine introduction studies and disease surveillance will be required to obtain necessary data to inform long-term impacts of interventions against enteric pathogens. As new enteric pathogen vaccines are being considered for introduction, robust population-level surveys of key outcomes like stunting and neurodevelopment will be critical to have in place so the full value of these vaccines can be informed by real data. The field can draw on other pathogen examples-such as measles which was studied post-vaccine introduction, to further informs the value proposition of this important vaccine(39).

This review had several limitations. To represent the breadth of ways in which enteric pathogens and longer-term outcomes are reported in the literature, we did not restrict studies no limit abstracted information to specific exposure and outcome framings leading to difficulty in interpretation across heterogeneous measures and comparison groups. We additionally allowed for multiple publications from the same underlying cohort to be included in the review when outcomes or comparison groups varied slightly. This approach further added to heterogeneity and difficulty in interpretation. Individual participant data meta-analysis could overcome these challenges, albeit with a significantly larger time and resource investment. Given the challenges with confounding and reverse causality, we do not believe that additional observational studies are needed. Instead, investment in pathogen-targeted randomized controlled trials, and specifically, post-introduction (phase 4) vaccine trials will likely be the optimal setting for estimation of true pathogen-specific associations with the most salient long-term of enteric pathogens, such as stunting and neurocognitive delay. Additionally, consensus on the most relevant outcomes to include, such as change in LAZ/HAZ between enrollment and 24 months of follow-up, will hasten future synthesis exercises.

## Data Availability

All data is available in the manuscript and associated tables.

## Contributions

MHA and BG conceptualized the review; GZ, MHA, ML, FA, PS developed the search criteria; ML, FA, GZ conducted the literature review with PP reconciling any disagreements in application of inclusion/exclusion criteria. All authors contributed to interpretation, writing and reviewing the manuscript.

## Ethical approval statement

Human subjects ethical approval was not required as no human subjects were involved in this research.

## References

1. Villavicencio F, Perin J, Eilerts-Spinelli H, Yeung D, Prieto-Merino D, Hug L, et al. Global, regional, and national causes of death in children and adolescents younger than 20 years: an open data portal with estimates for 2000-21. Lancet Glob Health. 2024;12(1):e16–e7.

2. Anderson JDt, Bagamian KH, Muhib F, Amaya MP, Laytner LA, Wierzba T, et al. Burden of enterotoxigenic Escherichia coli and shigella non-fatal diarrhoeal infections in 79 low-income and lower middle-income countries: a modelling analysis. Lancet Glob Health. 2019;7(3):e321–e30.

3. Brennhofer SA, Platts-Mills JA, Lewnard JA, Liu J, Houpt ER, Rogawski McQuade ET. Antibiotic use attributable to specific aetiologies of diarrhoea in children under 2 years of age in low-resource settings: a secondary analysis of the MAL-ED birth cohort. BMJ Open. 2022;12(4):e058740.

4. Hasso-Agopsowicz M, Lopman BA, Lanata CF, Rogawski McQuade ET, Kang G, Prudden HJ, et al. World Health Organization Expert Working Group: Recommendations for assessing morbidity associated with enteric pathogens. Vaccine. 2021;39(52):7521–5.

5. Baker JM, Hasso-Agopsowicz M, Pitzer VE, Platts-Mills JA, Peralta-Santos A, Troja C, et al. Association of enteropathogen detection with diarrhoea by age and high versus low child mortality settings: a systematic review and meta-analysis. Lancet Glob Health. 2021;9(10):e1402–e10.

6. Asare EO, Hergott D, Seiler J, Morgan B, Archer H, Wiyeh AB, et al. Case fatality risk of diarrhoeal pathogens: a systematic review and meta-analysis. Int J Epidemiol. 2022;51(5):1469–80.

7. Libby TE, Delawalla MLM, Al-Shimari F, MacLennan CA, Vannice KS, Pavlinac PB. Consequences of Shigella infection in young children: a systematic review. Int J Infect Dis. 2023;129:78–95.

8. von Elm E, Altman DG, Egger M, Pocock SJ, Gøtzsche PC, Vandenbroucke JP. The Strengthening the Reporting of Observational Studies in Epidemiology (STROBE) statement: guidelines for reporting observational studies. J Clin Epidemiol. 2008;61(4):344–9.

9. Investigators M-EN. The MAL-ED study: a multinational and multidisciplinary approach to understand the relationship between enteric pathogens, malnutrition, gut physiology, physical growth, cognitive development, and immune responses in infants and children up to 2 years of age in resource-poor environments. Clin Infect Dis. 2014;59 Suppl 4:S193–206.

10. Kotloff KL, Blackwelder WC, Nasrin D, Nataro JP, Farag TH, van Eijk A, et al. The Global Enteric Multicenter Study (GEMS) of diarrheal disease in infants and young children in developing countries: epidemiologic and clinical methods of the case/control study. Clinical infectious diseases : an official publication of the Infectious Diseases Society of America. 2012;55 Suppl 4:S232–45.

11. Kirkpatrick BD, Colgate ER, Mychaleckyj JC, Haque R, Dickson DM, Carmolli MP, et al. The "Performance of Rotavirus and Oral Polio Vaccines in Developing Countries" (PROVIDE) study: description of methods of an interventional study designed to explore complex biologic problems. Am J Trop Med Hyg. 2015;92(4):744–51.

12. Black RE, Brown KH, Becker S. Effects of diarrhea associated with specific enteropathogens on the growth of children in rural Bangladesh. Pediatrics. 1984;73(6):799–805.

13. Donowitz JR, Drew J, Taniuchi M, Platts-Mills JA, Alam M, Ferdous T, et al. Diarrheal Pathogens Associated With Growth and Neurodevelopment. Clin Infect Dis. 2021;73(3):e683–e91.

14. Nasrin D, Blackwelder WC, Sommerfelt H, Wu Y, Farag TH, Panchalingam S, et al. Pathogens Associated With Linear Growth Faltering in Children With Diarrhea and Impact of Antibiotic Treatment: The Global Enteric Multicenter Study. J Infect Dis. 2021;224(12 Suppl 2):S848–s55.

15. Rogawski ET, Liu J, Platts-Mills JA, Kabir F, Lertsethtakarn P, Siguas M, et al. Use of quantitative molecular diagnostic methods to investigate the effect of enteropathogen infections on linear growth in children in low-resource settings: longitudinal analysis of results from the MAL-ED cohort study. Lancet Glob Health. 2018;6(12):e1319–e28.

16. Relationship between growth and illness, enteropathogens and dietary intakes in the first 2 years of life: findings from the MAL-ED birth cohort study. BMJ Glob Health. 2017;2(4):e000370.

17. Haque MA, Nasrin S, Palit P, Das R, Wahid BZ, Gazi MA, et al. Site-Specific Analysis of the Incidence Rate of Enterotoxigenic Escherichia coli Infection Elucidates an Association with Childhood Stunting, Wasting, and Being Underweight: A Secondary Analysis of the MAL-ED Birth Cohort. Am J Trop Med Hyg. 2023;108(6):1192–200.

18. Luoma J, Adubra L, Ashorn P, Ashorn U, Bendabenda J, Dewey KG, et al. Association between asymptomatic infections and linear growth in 18-24-month-old Malawian children. Matern Child Nutr. 2023;19(1):e13417.

19. Palit P, Das R, Haque MA, Hasan MM, Noor Z, Mahfuz M, et al. Risk Factors for Norovirus Infections and Their Association with Childhood Growth: Findings from a Multi-Country Birth Cohort Study. Viruses. 2022;14(3).

20. Lee G, Paredes Olortegui M, Peñataro Yori P, Black RE, Caulfield L, Banda Chavez C, et al. Effects of Shigella-, Campylobacter- and ETEC-associated diarrhea on childhood growth. Pediatr Infect Dis J. 2014;33(10):1004–9.

21. Hossain MI, Nasrin S, Das R, Palit P, Sultana AA, Sobi RA, et al. Symptomatic and Asymptomatic Campylobacter Infections and Child Growth in South Asia: Analyzing Data from the Global Enteric Multicenter Study. Am J Trop Med Hyg. 2023;108(6):1204–11.

22. Platts-Mills JA, Gratz J, Mduma E, Svensen E, Amour C, Liu J, et al. Association between stool enteropathogen quantity and disease in Tanzanian children using TaqMan array cards: a nested case-control study. Am J Trop Med Hyg. 2014;90(1):133–8.

23. Rogawski ET, Liu J, Platts-Mills JA, Kabir F, Lertsethtakarn P, Siguas M, et al. Use of quantitative molecular diagnostic methods to investigate the effect of enteropathogen infections on linear growth in children in low-resource settings: longitudinal analysis of results from the MAL-ED cohort study. Lancet Glob Health. 2018;6(12):e1319–e28.

24. Rouhani S, Griffin NW, Yori PP, Olortegui MP, Siguas Salas M, Rengifo Trigoso D, et al. Gut Microbiota Features Associated With Campylobacter Burden and Postnatal Linear Growth Deficits in a Peruvian Birth Cohort. Clin Infect Dis. 2020;71(4):1000–7.

25. Haque MA, Platts-Mills JA, Mduma E, Bodhidatta L, Bessong P, Shakoor S, et al. Determinants of Campylobacter infection and association with growth and enteric inflammation in children under 2 years of age in low-resource settings. Sci Rep. 2019;9(1):17124.

26. Amour C, Gratz J, Mduma E, Svensen E, Rogawski ET, McGrath M, et al. Epidemiology and Impact of Campylobacter Infection in Children in 8 Low-Resource Settings: Results From the MAL-ED Study. Clin Infect Dis. 2016;63(9):1171–9.

27. Kabir F, Iqbal J, Jamil Z, Iqbal NT, Mallawaarachchi I, Aziz F, et al. Impact of enteropathogens on faltering growth in a resource-limited setting. Front Nutr. 2022;9:1081833.

28. Das R, Haque MA, Chisti MJ, Faruque ASG, Ahmed T. Associated factors, post infection child growth, and household cost of invasive enteritis among under 5 children in Bangladesh. Sci Rep. 2021;11(1):12738.

29. Schnee AE, Haque R, Taniuchi M, Uddin MJ, Alam MM, Liu J, et al. Identification of Etiology-Specific Diarrhea Associated With Linear Growth Faltering in Bangladeshi Infants. Am J Epidemiol. 2018;187(10):2210–8.

30. Scharf RJ, McQuade ETR, Svensen E, Huggins A, Maphula A, Bayo E, et al. Early-Life Enteric Pathogen Exposure, Socioeconomic Status, and School-Age Cognitive Outcomes. Am J Trop Med Hyg. 2023;109(2):436–42.

31. Das R, Palit P, Haque MA, Ahmed T, Faruque ASG. Association between Pathogenic Variants of Diarrheagenic Escherichia coli and Growth in Children under 5 Years of Age in the Global Enteric Multicenter Study. Am J Trop Med Hyg. 2022;107(1):72–81.

32. George CM, Burrowes V, Perin J, Oldja L, Biswas S, Sack D, et al. Enteric Infections in Young Children are Associated with Environmental Enteropathy and Impaired Growth. Trop Med Int Health. 2018;23(1):26–33.

33. Platts-Mills JA, Taniuchi M, Uddin MJ, Sobuz SU, Mahfuz M, Gaffar SA, et al. Association between enteropathogens and malnutrition in children aged 6-23 mo in Bangladesh: a case-control study. Am J Clin Nutr. 2017;105(5):1132–8.

34. Das R, Haque MA, Kotloff KL, Nasrin D, Hossain MJ, Sur D, et al. Enteric viral pathogens and child growth among under-five children: findings from South Asia and sub-Saharan Africa. Sci Rep. 2024;14(1):13871.

35. González-Fernández D, Cousens S, Rizvi A, Chauhadry I, Soofi SB, Bhutta ZA. Infections and nutrient deficiencies during infancy predict impaired growth at 5 years: Findings from the MAL-ED study in Pakistan. Front Nutr. 2023;10:1104654.

36. Sanchez JJ, Alam MA, Stride CB, Haque MA, Das S, Mahfuz M, et al. Campylobacter infection and household factors are associated with childhood growth in urban Bangladesh: An analysis of the MAL-ED study. PLoS Negl Trop Dis. 2020;14(5):e0008328.

37. Pavlinac PB, Platts-Mills J, Liu J, Atlas HE, Gratz J, Operario D, et al. Azithromycin for bacterial watery diarrhea: A reanalysis of the AntiBiotics for Children with severe Diarrhea (ABCD) trial incorporating molecular diagnostics. J Infect Dis. 2023.

38. Pinkerton R, Oria RB, Lima AA, Rogawski ET, Oria MO, Patrick PD, et al. Early Childhood Diarrhea Predicts Cognitive Delays in Later Childhood Independently of Malnutrition. Am J Trop Med Hyg. 2016;95(5):1004–10.

39. Aaby P, Samb B, Simondon F, Seck AM, Knudsen K, Whittle H. Non-specific beneficial effect of measles immunisation: analysis of mortality studies from developing countries. BMJ. 1995;311(7003):481–5.

