## Supplemental Files for "Anthropometric and neurocognitive consequences of *Campylobacter*, enterotoxigenic *Escherichia coli*, and norovirus: A systematic review"

**Supplementary Material**

Table S1. Search terms

| **Sequelae terms** | ("Learning Disabilities"[Mesh] OR "Cognition Disorders"[Mesh:NoExp] OR "Cognitive Dysfunction"[Mesh] OR "Learning Disabilit*"[tiab] OR “Cognitive Development”[tiab] OR “Cogniti*”[tiab] OR "Child Development"[Mesh] OR "Growth Disorders"[Mesh] OR "Body Size"[Mesh] OR "child development" OR "postnatal development" OR "post-natal development" OR growth[tiab] OR "Crown Rump Length" OR height OR stunting OR stunted) |
| --- | --- |
| **Pathogens and disease presentation** | AND ("Campylobacter"[Mesh] OR "Campylobacter Infections"[Mesh] OR Campylobacter*[tiab] OR "Norovirus"[Mesh] OR Norovirus*[tiab] OR "Enterotoxigenic Escherichia coli"[Mesh] OR ETEC[tiab] OR "Enterotoxigenic E*"[tiab] OR "Diarrhea"[Mesh] OR "Dysentery"[Mesh:NoExp]) |
| **Age group terms:** | AND ("Child, Preschool"[Mesh] OR "Infant"[Mesh] OR "Child*"[tw] OR "Infant*"[All Fields] OR "Newborn"[All Fields] OR "Baby"[All Fields] OR "Babies"[All Fields] OR "Neonat*"[All Fields] OR "Pediatric"[tw] OR "Paediatric"[tw]) |
| **Publication date** | AND “1980/01/01”[Date - Publication] : “2024/21/08 [Date - Publication] |
| **Publication type** | NOT (“Editorial”[Publication Type] OR “Letter”[Publication Type] OR “Review”[Publication Type] OR “Case Reports”[Publication Type]) |

Table S2. Quality reporting scale utilized for studies included in systematic review*

| **Method head** | **Description** | **Score** |
| --- | --- | --- |
| **STROBE Guidelines** | |  |
| **Study design** | Present key elements of study design early in the paper | 1 |
| **Setting** | Describe the setting, locations, and relevant dates, including periods of recruitment, exposure, follow-up, and data collection | 1 |
| **Participants** | (*a*) *Cohort study*—Give the eligibility criteria, and the sources and methods of selection of participants. Describe methods of follow-up  *Case-control study*—Give the eligibility criteria, and the sources and methods of case ascertainment and control selection. Give the rationale for the choice of cases and controls  *Cross-sectional study*—Give the eligibility criteria, and the sources and methods of selection of participants | 1 |
|  | (*b*) *Cohort study*—For matched studies, give matching criteria and number of exposed and unexposed  *Case-control study*—For matched studies, give matching criteria and the number of controls per case |  |
| **Variables** | Clearly define all outcomes, exposures, predictors, potential confounders, and effect modifiers. Give diagnostic criteria, if applicable | 1 |
| **Data sources/** **measurement** | For each variable of interest, give sources of data and details of methods of assessment (measurement). Describe comparability of assessment methods if there is more than one group | 1 |
| **Bias** | Describe any efforts to address potential sources of bias | 1 |
| **Study size** | Explain how the study size was arrived at | 1 |
| **Quantitative** **variables** | Explain how quantitative variables were handled in the analyses. If applicable, describe which groupings were chosen and why | 1 |
| **Statistical** **methods** | (*a*) Describe all statistical methods, including those used to control for confounding | 1 |
|  | (*b*) Describe any methods used to examine subgroups and interactions |  |
|  | (*c*) Explain how missing data were addressed |  |
|  | (*d*) *Cohort study*—If applicable, explain how loss to follow-up was addressed  *Case-control study*—If applicable, explain how matching of cases and controls was addressed  *Cross-sectional study*—If applicable, describe analytical methods taking account of sampling strategy | 1 |
|  | (*e*) Describe any sensitivity analyses |  |
| **Adapted from: von Elm E, Altman DG, Egger M, Pocock SJ, Gøtzsche PC, Vandenbroucke JP. The Strengthening the Reporting of Observational Studies in Epidemiology (STROBE) statement: guidelines for reporting observational studies. J Clin Epidemiol 2008;* ***61****(4): 344-9.* | | |

Table S3. Quality ratings of included studies

| **Study** | **Country** | **Recruitment Years** | **Study name (if applicable)** | **Adapted STROBE score** |
| --- | --- | --- | --- | --- |
| Amour 2016 | Bangladesh, India, Nepal, South Africa, Tanzania, Brazil, Peru | 2009-2012 | MAL-ED | 8 |
| Black 1984 | Bangladesh | 1978-1979 | Not named | 8 |
| Bray 2019 | Bangladesh | 2007-2010 | GEMS | 5 |
| Caulfield 2017 | Bangladesh, India, Nepal, South Africa, Tanzania, Brazil, Peru | 2009-2014 | MAL-ED | 9 |
| Das 2021 | Bangladesh | 2007-2011 | GEMS | 7 |
| Das 2022 | Bangladesh, India, Pakistan, The Gambia, Mali, Mozambique,  Kenya | 2007-2011 | GEMS | 8 |
| Das 2024 | Bangladesh, India, Pakistan, The Gambia, Mali, Mozambique,  Kenya | 2007-2011 | GEMS | 8 |
| Diaz 2023 | Haiti | 2020-2021 |  | 8 |
| Donowitz 2021 | Bangladesh | 2014-2016 |  | 6 |
| George 2017 | Bangladesh | 2014 |  | 6 |
| George 2023 | Democratic Republic of the Congo | 2018-2019 | REDUCE | 7 |
| González-Fernández 2023 | Pakistan | 2010-2012 | MAL-ED | 8 |
| Haque 2019 | Bangladesh, India, Nepal, South Africa, Tanzania, | 2009-2012 | MAL-ED | 8 |
| Haque 2023 | Bangladesh, Brazil, India, Nepal, Peru, Pakistan, South Africa, Tanzania | 2009-2012 | MAL-ED | 9 |
| Hossain 2023 | Bangladesh, India, Pakistan | 2007-2011 | GEMS | 8 |
| Iqbal 2019 | Pakistan | 2012-2015 |  | 6 |
| Kabir 2022 | Pakistan | 2016-2018 | SEEM | 6 |
| Lee 2013 | Peru | 2002-2006 | Not named but same cohort as Lee 2014 | 8 |
| Lee 2014 | Peru | 2002-2006 | Not named but same cohort as Lee 2013 | 8 |
| Luoma 2023 | Malawi | 2009-2011 | iLiNS‐DYAD‐M | 9 |
| Nasrin 2021 | The Gambia, Mali, Mozambique, Kenya, Pakistan, Bangladesh, India | 2007-2011 | GEMS | 10 |
| Pajuelo 2024 | Peru | 2016-2019 |  | 7 |
| Palit 2022 | Bangladesh, India, Nepal, South Africa, Tanzania, Brazil, Peru |  | MAL-ED | 8 |
| Platts-Mills 2014 | Tanzania | 2009-2012 | MAL-ED | 7 |
| Platts-Mills 2017 | Bangladesh | 2009-2012 | PROVIDE | 7 |
| Rogawski 2018 | Bangladesh, India, Nepal, South Africa, Tanzania, Brazil, Peru | 2009-2012 | MAL-ED | 9 |
| Rouhani 2020 | Peru | 2009-2012 | MAL-ED | 8 |
| Sanchez 2020 | Bangladesh | 2010-2012 | MAL-ED | 8 |
| Scharf 2023 | Brazil, Tanzania, South Africa | 2009-2012 | MAL-ED | 8 |
| Schnee 2018 | Bangladesh | 2011-2014 | PROVIDE | 8 |
